# Polygenic analyses show important differences between MDD symptoms collected using PHQ9 and CIDI-SF

**DOI:** 10.1101/2023.02.27.23286527

**Authors:** Lianyun Huang, Sonja Tang, Jolien Rietkerk, Vivek Appadurai, Morten Dybdahl Krebs, Andrew J. Schork, Thomas Werge, Verena Zuber, Kenneth Kendler, Na Cai

## Abstract

Symptoms of Major Depressive Disorder (MDD) are commonly assessed using self-rating instruments like the Patient Health Questionnaire 9 (PHQ9, for current symptoms), and the Composite International Diagnostic Interview Short-Form (CIDI-SF, for lifetime worst-episode symptoms). Using data from the UKBiobank, we show that corresponding symptoms endorsed through PHQ9 and CIDI-SF have low to moderate genetic correlations (rG=0.43-0.87), and this cannot be fully attributed to different severity thresholds or the use of a skip-structure in CIDI-SF. Through a combination of Mendelian Randomization (MR) and polygenic prediction analyses, we find that PHQ9 symptoms are more associated with traits which reflect general dysphoria, while the skip-structure in CIDI-SF allows for the identification of heterogeneity among likely MDD cases. This has important implications on factor analyses performed on their respective genetic covariance matrices for the purpose of identification of genetic factors behind MDD symptom dimensions and heterogeneity.

## Introduction

Two sets of symptom-level data of Major Depressive Disorder (MDD) are available in the UKBiobank^1^ through the self-administered online Mental Health Questionnaire (MHQ)^2^. First, current symptoms of MDD are assessed through the Patient Health Questionnaire 9 (PHQ9)^3^, a screening tool that scores the occurrence of all 9 DSM5-based symptoms for MDD^4^ in the past two weeks. A high sum score is used as an indicator of a potential presence of MDD and the basis for recommending clinical assessment. Second, MDD symptoms experienced during the lifetime worst episode of MDD are assessed through the Composite International Diagnostic Interview Short Form (CIDI-SF)^5,6^, which contains a “skip-structure” : 7 out of 9 MDD symptoms are only assessed when two weeks of sad mood or anhedonia are endorsed (cardinal symptoms of MDD, **Figure 1A**). As a result, the CIDI-SF assesses WorstEpisode symptoms in a clinically enriched population, which has a smaller sample size (∼50K, **Figure 1B**) than the PHQ9 symptoms assessed in the general population(∼100K).

**Figure 1:**
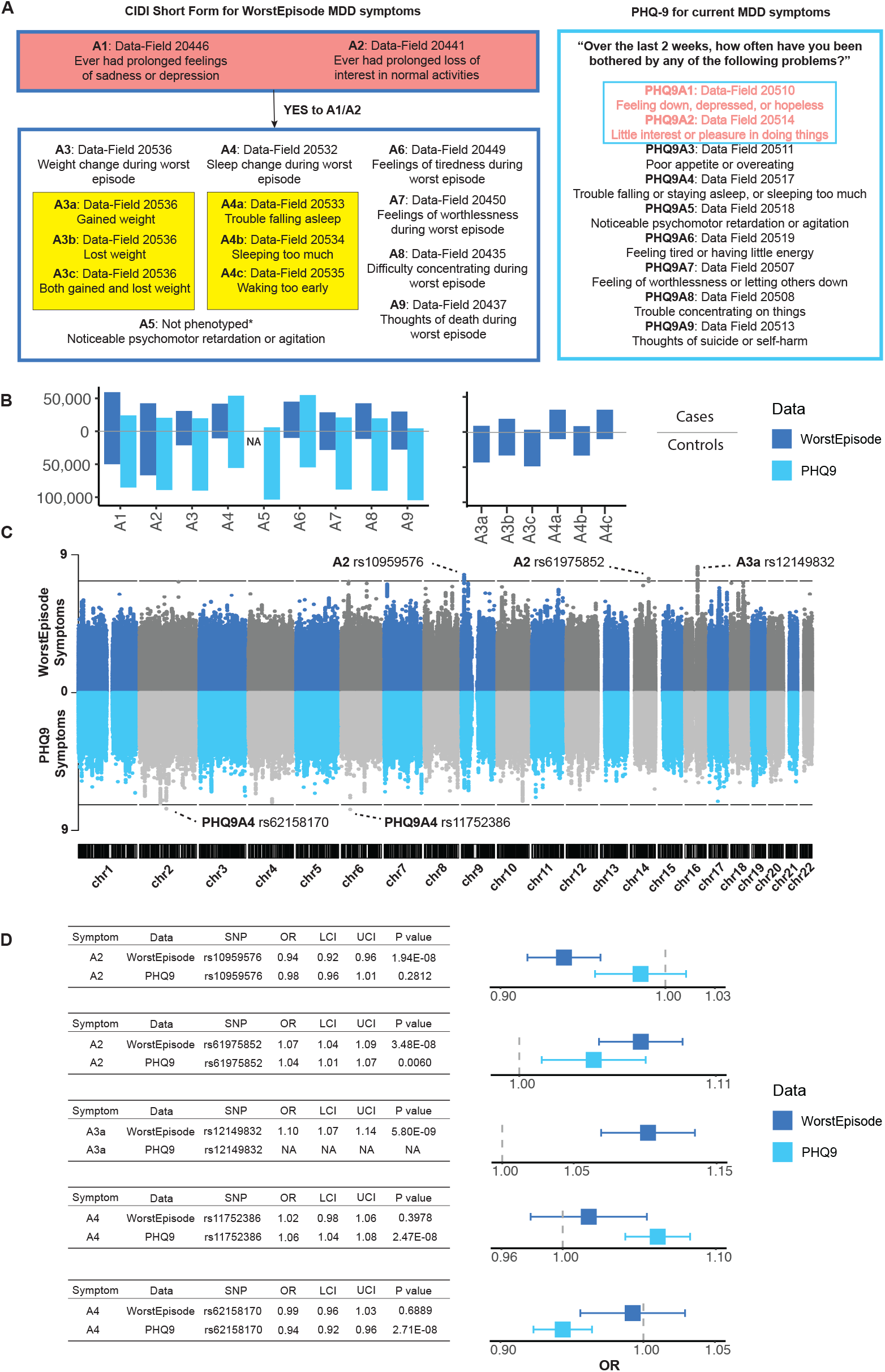
Definition, sample size and GWAS of WorstEpisode and PHQ9 symptoms of MDD in UK Biobank. **A**. Definitions of WorstEpisode and PHQ9 symptoms of MDD in UK Biobank. **B**. Sample sizes in WorstEpisode and PHQ9 symptoms. There is no data for WorstEpisode A5; subgroups of symptoms A3 and A4 are not assessed in PHQ9. **C**. Miami plot for 14 WorstEpisode symptoms on top and 9 PHQ9 symptoms at the bottom. Associations with P-values smaller than 5×10^−8^ are considered as genome-wide significant and are indicated in the plot. **D**. Forest plots and accompanying data showing the odds ratios (OR) and P-values at significant loci; statistics at the corresponding WorstEpisode or PHQ9 symptoms are shown for comparison; error bars show 95% confidence intervals of the OR estimates.

Previous studies have performed genome-wide association studies (GWAS) on individual items in the PHQ9^7^. Likewise, genetic covariance-based factor analyses using genomicSEM^8^ have been performed on PHQ9 symptoms to identify symptom dimensions of MDD and how they overlap with those of anxiety and neuroticism^7,9^. In contrast, there are no genetic studies that analyse lifetime WorstEpisode symptoms or compare them with PHQ9 symptoms. This is despite the much wider use of WorstEpisode symptoms in phenotypic covariance-based factor analysis over the past decades^10– 12^, and findings that suggest symptoms in clinically-enriched populations have different structures and meanings from those measured in a general population^10^.

In this paper, we ask whether PHQ9 symptoms capture the same underlying biology as WorstEpisode symptoms, using data collected through the MHQ in UKBiobank^1^. In particular, we want to know if the two sets of symptoms may be interchangeably used or combined to study symptom-level genetic risks and MDD heterogeneity. We find that while PHQ9 and WorstEpisode symptoms have similar liability scale SNP-heritabilities (h^2^_SNP_), they have distinct genetic components. Overall, PHQ9 symptoms have greater genetic sharing with subjective well-being, insomnia, neuroticism, anxiety, and exposure to stressful life events. Polygenic predictions on MDD and 50 non-MDD phenotypes show a clear distinction between symptoms assessed with both questionnaires due to the skip-structure inherent in CIDI-SF; it enforces those symptoms assessed conditional on cardinal symptom endorsement to capture genetic sources of heterogeneity between likely MDD cases, rather than genetic liability to MDD. Finally, factor analyses performed on genetic covariance from both sets of symptoms using genomicSEM suggest different structures, with WorstEpisode evidencing factor structures concordant with previous factor analyses using phenotypic covariance from WorstEpisode symptoms in clinical cohorts.

We, therefore, conclude that PHQ9 and WorstEpisode symptoms do not reflect the same biology, and are therefore not interchangeable in analysis - the former indexes genetic liability to general dysphoria in addition to MDD, while the latter captures genetic heterogeneity among likely MDD cases.

## Results

### GWAS on PHQ9 and WorstEpisode symptoms

We first perform GWAS (**Methods**) on 14 WorstEpisode symptoms (**Figure 1C, Methods, Supplementary Table 1**) and 9 PHQ9 symptoms (**Figure 1C, Methods, Supplementary Table 2**). We find a total of 3 significantly associated loci (at P-value < 5×10^−8^) in 2 out of the 14 WorstEpisode symptoms (**Figure 1C, Supplementary Table 3**): rs10959576 (OR = 0.94; 95% CI = [0.92-0.96], P-value = 1.94×10^−8^) and rs61975852 (OR = 1.07; 95% CI = [1.04-1.09], P-value = 3.48×10^−8^) for symptom A2 (anhedonia), and rs12149832 (OR = 1.10; 95% CI = [1.07-1.13], P-value = 5.80×10^−9^) for symptom A3a (increase in appetite or weight). We find 2 significantly associated loci in 1 out of 9 PHQ9 symptoms (**Figure 1C, Supplementary Table 3**): rs11752386 (OR = 1.06; 95% CI = [1.04-1.09], P-value = 2.47×10^−8^) and rs62158170 (OR = 0.94; 95% CI = [0.94-0.96], P-value = 2.71×10^−8^) for A4 (change in sleep). None are significant once we correct for multiple testing on the number of symptoms analysed.

We ask if each significant locus for WorstEpisode symptoms has a similar effect on the corresponding PHQ9 symptom, and vice versa. We find that only one of the SNPs significantly associated with WorstEpisode A2 has a significant association (P < 0.05/5) in the same direction of effect for A2 assessed through PHQ9 (rs61975852, P-value = 0.006, **Figure 1D, Supplementary Table 3**). As GWAS power is different between the two sets of symptoms due to differences in sample sizes (**Figure 1B**), we calculate the effective sample size (N_eff_) of each corresponding pair of PHQ9 and WorstEpisode symptoms, and down-sample the larger of the two to the lower N_eff_, keeping prevalence constant (**Methods, Supplementary Table 1, 2**). We find that though none of the significant loci remain significant in GWAS after down-sampling, their relative effect sizes in PHQ9 and WorstEpisode symptoms remain the same (**Supplementary Figure 1**).

### Genetic differences between PHQ9 and WorstEpisode symptoms are not fully due to skip-structure or severity thresholds

We next estimate liability scale SNP-heritability (h^2^_SNP_) for all WorstEpisode and PHQ9 symptoms using LD score regression (LDSC^13^, **Methods**). We find that while there are differences in h^2^_SNP_ estimates between the corresponding PHQ9 and WorstEpisode symptoms, their error bars overlap in all instances (**Figure 2A**). The genetic correlations (rG)^14^ between corresponding WorstEpisode and current symptoms, however, are mostly significantly different from unity, with the exception of A3 (change in appetite) and A9 (suicide ideation, **Figure 2B**), demonstrating that WorstEpisode and PHQ9 symptoms are driven by partly distinct genetic factors.

**Figure 2:**
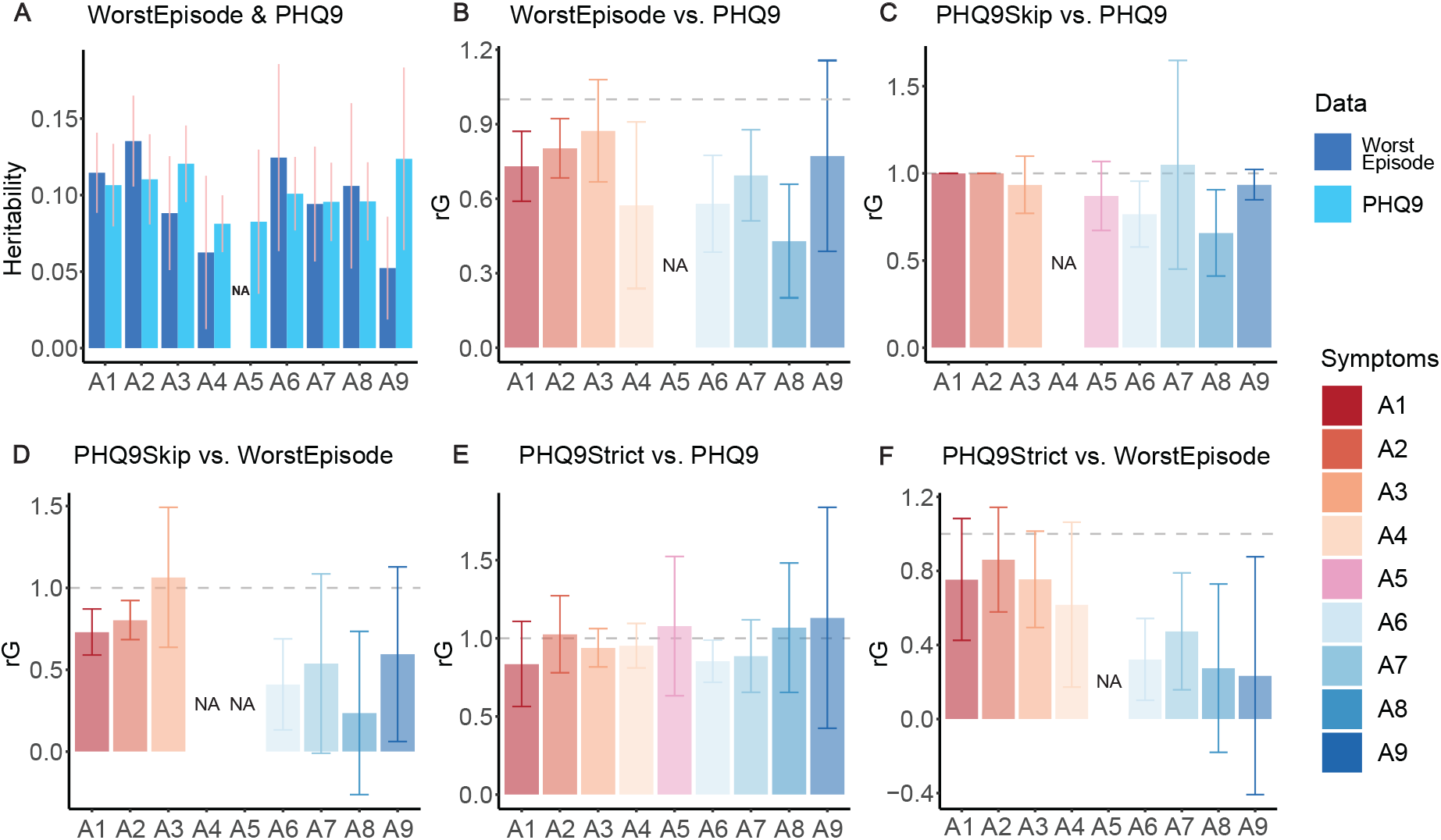
SNP heritability and genetic correlation estimates from LDSC. **A**. Liability scale h^2^_SNP_ estimates for each WorstEpisode and PHQ9 symptom, calculated assuming their observed prevalence in UKBiobank are equal to their population prevalence. **B**. Genetic correlation (rG) between WorstEpisode and PHQ9 symptoms. **C**. rG between PHQ9 and PHQ9Skip symptoms. **D**. rG between PHQ9Skip and WorstEpisode symptoms. **E**. rG between PHQ9Strict and PHQ9 symptoms. **F**. rG between PHQ9Strict and WorstEpisode symptoms. For all plots, horizontal dashed lines show rG = 1; error bars indicate 95% confidence intervals of estimates; lack of convergence on the estimate or missing data is indicated as “NA”.

As WorstEpisode symptoms are assessed with a skip-structure in the CIDI-SF, and the PHQ9 symptoms are not, we ask if their low rG is due to this inherent difference in the instruments. To do this, we implement the skip-structure on PHQ9 symptoms, like in CIDI-SF, obtaining “PHQ9Skip” symptoms (**Supplementary Methods, Supplementary Table 2**). We find that their rGs with PHQ9 symptoms are not significantly different from unity (**Figure 2C**), whereas their rGs with WorstEpisode symptoms remain low (**Figure 2D**), similar to those between WorstEpisodes and PHQ9 symptoms (**Figure 2B**). In other words, low rG between PHQ9 and WorstEpisode symptoms cannot be largely attributed to the skip-structure in WorstEpisode assessments.

We then ask if the low rGs are due to different severity thresholds in the two sets of symptoms. As shown in **Figure 1A** and defined in previous studies ^7,9^, any occurrence of a symptom in PHQ9 would qualify as an endorsement (**Supplementary Table 2**). In comparison, we require an occurrence of “nearly every day” to qualify as an endorsement of WorstEpisode symptom. We, therefore, define a set of “PHQ9Strict” symptoms, requiring the frequency of symptoms to be “nearly every day” (**Supplementary Methods, Supplementary Table 2**). Like PHQ9Skip symptoms shown above, PHQ9Strict symptoms show similar h^2^_SNP_ to PHQ9 symptoms, and rGs between them are not significantly different from unity (**Figure 2E**). Further, just like PHQ9 symptoms, PHQ9Strict symptoms show low rG with WorstEpisode symptoms (**Figure 2F**). This shows that the occurrence of PHQ9 symptoms is indicative of severity; lowering the occurrence threshold does not introduce heterogeneity in genetic contributions, and does not account for the difference in the genetic contribution to PHQ9 and WorstEpisode symptoms. Overall, our findings suggest that the low genetic correlations between the PHQ9 and WorstEpisode depressive symptoms cannot be fully explained by methodological differences in how they are assessed.

### Endorsement of PHQ9 symptoms is more likely due to general dysphoria

It is well known that long-standing conditions that cause general dysphoria can lead to inflation in self-ratings of current symptoms with PHQ9^15,16^. To test this, we first ask if four traits including insomnia and measures of subjective well-being have greater genetic sharing with endorsements of the most similarly phrased PHQ9 items than the corresponding WorstEpisode symptom in CIDI-SF (**Supplementary Table 4**). We find all four traits have higher rG with PHQ9 symptoms than WorstEpisode symptoms, even though their error bars overlap (one-sided paired t-test P = 0.02, **Figure 3A**). We then perform the same analysis with four further sets of phenotypes: anxiety symptoms from the GAD7 questionnaire in the MHQ^17,18^; experience of stressful life events both recently and in one’s lifetime^19–21^, and individual items of the neuroticism items in the Eysenck Personality Questionnaire Revised-Short Form (EPQR-S)^22^ (**Methods, Supplementary Table 4**). Consistently, PHQ9 symptoms have higher rG with all four sets of phenotypes than WorstEpisode symptoms (one-sided paired t-test P for neuroticism items = 2.20×10^−16^, anxiety symptoms = 2.93×10^− 11^; lifetime trauma = 2.16×10^−7^, recent stressful life events = 0.0002, **Figure 3B**). Overall, we find that PHQ9 symptoms have greater genetic sharing with these non-MDD phenotypes that index general dysphoria.

**Figure 3:**
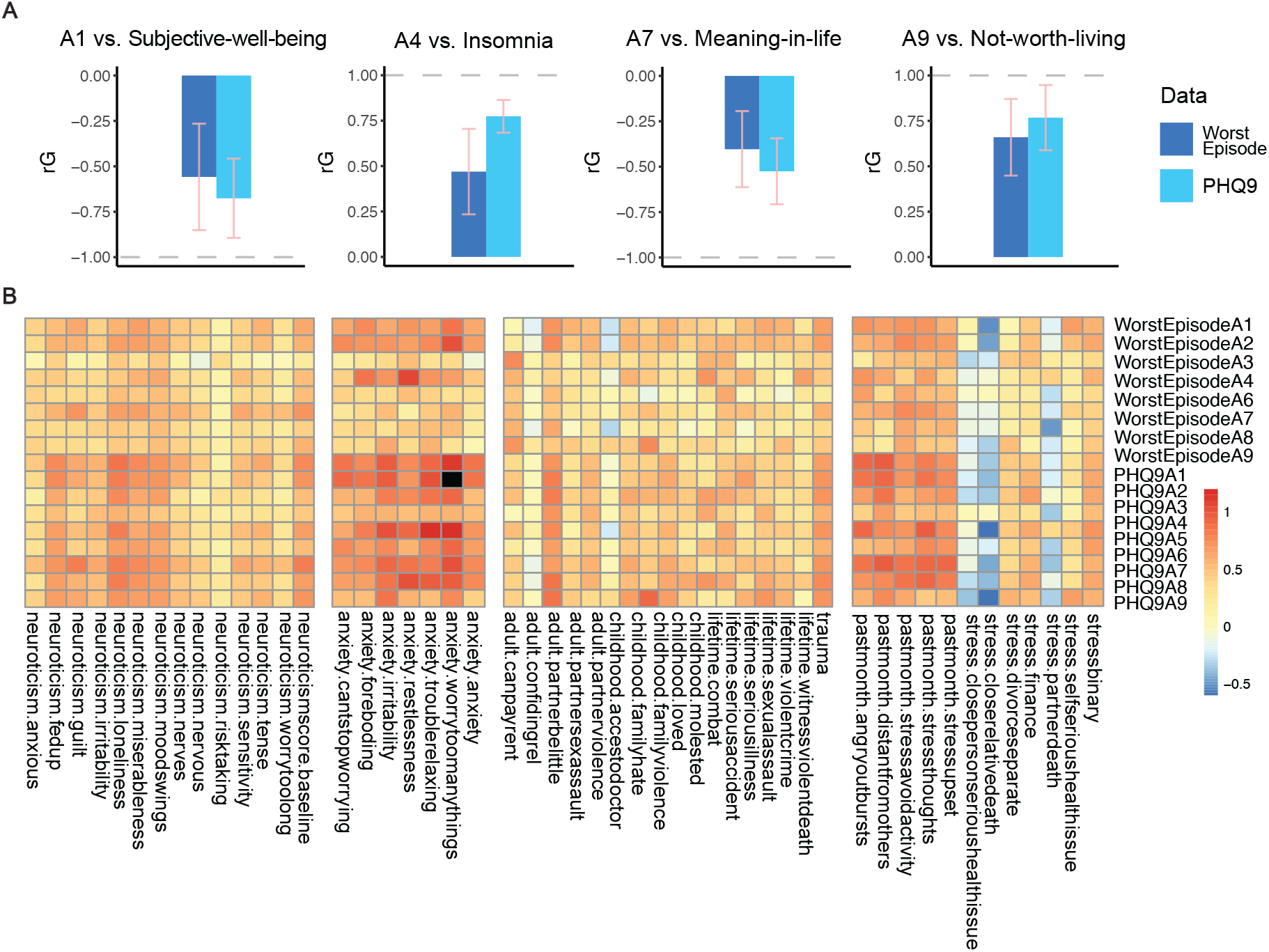
Genetic correlation (rG) estimates between PHQ9 or WorstEpisode symptoms and non-MDD phenotypes. **A**. rG between depressed mood (A1) vs. subjective well-being; change of sleep (A4) vs. insomnia; feeling of worthlessness (A7) vs. finding meaning in life; and suicidal ideation (A9) vs. finding life not worth living, in both WorstEpisode and PHQ9 symptom definitions. Error bars indicate 95% confidence intervals of the estimates. Bar directions indicate either positive or negative rG. **B**. rG between PHQ9 or WorstEpisode symptoms of MDD vs. neuroticism items in the EPQR-S^22^; anxiety symptoms from the GAD7 questionnaire in the MHQ^17,18^; and experience of stressful life events both recently and in one’s lifetime^19–21^; black square indicates a lack of convergence on the estimate in LDSC.

We then ask if PHQ9 and WorstEpisode symptom endorsement may be partly due to general dysphoria, using Mendelian Randomization (MR), which assesses the association of genetic predictors of an exposure with an outcome^23,24^. Using univariable MR^25^ (UVMR, **Methods, Figure 4A**), we find that genetic effects on insomnia and measures of subjective well-being are associated with individual PHQ9 symptoms with a higher odds ratio than WorstEpisode symptoms (**Supplementary Figure 2, Supplementary Table 5**). Similarly, genetic effects on neuroticism score and generalised anxiety disorder (GAD) are all associated with all PHQ9 symptoms with significantly larger odds ratios than WorstEpisode symptoms (paired single-sided t-test P-value for neuroticism = 0.0013; anxiety = 0.0361, **Figure 4C-F, Supplementary Table 6**). This difference remains when both sets of symptoms are down-sampled to the same N_eff_ (**Supplementary Figure 3, Supplementary Table 6**). For recent stress and lifetime trauma, differences between the two sets of symptoms are not significant (**Supplementary Figure 3, Supplementary Table 6**).

**Figure 4:**
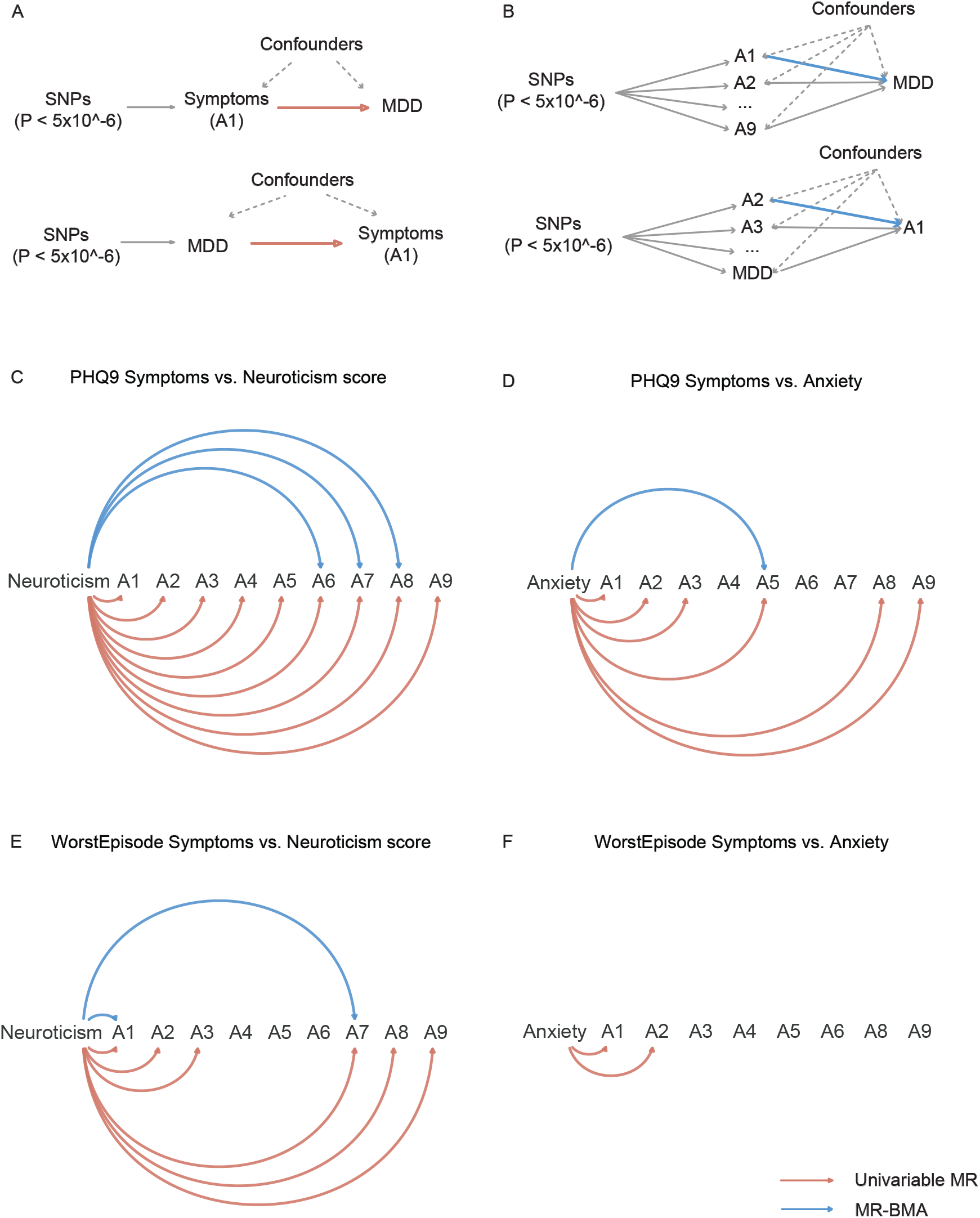
Mendelian randomization between MDD symptoms and non-MDD phenotypes. **A**. Directed Acyclic Graph (DAG) demonstrating the UVMR model between MDD symptoms and non-MDD phenotypes. **B**. DAG demonstrating the MR-BMA model between MDD symptoms and non-MDD phenotypes. **C-F**. UVMR results between neuroticism or anxiety (exposures) and PHQ9 or WorstEpisode symptoms (outcomes); orange arrows indicate significant UVMR results (P<0.05 after Bonferroni correction), blue arrows indicate significant MR-BMA results (<0.05 FDR). Marginal inclusion threshold for MR-BMA is >0.5.

Using a multivariable MR approach based on Bayesian model averaging^26^ (MR-BMA, **Figure 4B, Methods**), we further ask if genetic effects on neuroticism, anxiety, recent stress and lifetime trauma are associated with the endorsement of specific symptoms independently. For both WorstEpisode and PHQ9 symptoms, few UVMR associations remain significant in MR-BMA (**Figure 4C-F, Supplementary Figure 3, Supplementary Table 7**). In other words, general dysphoria contributes more to the endorsement of PHQ9 symptoms than WorstEpisode symptoms, though its contribution is non-specific in both symptom sets.

Finally, we ask if episodic MDD leads to the endorsement of either set of symptoms, and if either set of symptoms leads to episodic MDD. To do this, we perform two-sample UVMR and MR-BMA on either set of symptoms assessed in the UKBiobank with MDD assessed in external cohorts (PGC29^27^, iPSYCH2012^28^ and iPSYCH2015i^29^, **Methods**). To avoid overfitting, we do not perform this analysis using LifetimeMDD^30^ in UKBiobank, as it is defined by a combination of PHQ9 and WorstEpisode symptoms. We find that while genetic effects on MDD in all three cohorts show no significant associations on either set of symptoms, their effects are larger on PHQ9 symptoms across all three cohorts (paired single-sided t-test P-value for UVMR = 0.0004, **Supplementary Figure 4, Supplementary Table 8**). Conversely, while none of the genetic effects on PHQ9 symptoms are associated with MDD in all three cohorts, genetic effects on WorstEpisode symptoms A1, A2 and A7 are significantly associated with MDD in at least one cohort in either UVMR or MR-MBA analysis (**Supplementary Figure 4, Supplementary Table 8, 9**). All results still hold when comparing down-sampled PHQ9 and WorstEpisode symptoms (**Supplementary Figure 4, Supplementary Table 8, 9**). Overall, we find that genetic effects on WorstEpisode symptoms are associated with episodic MDD, while genetic effects on the latter are more associated with PHQ9 symptoms.

For all MR analyses, we verify that the genetic instruments used are strong (**Supplementary Methods, Supplementary Tables 5-9**). Further, all UVMR effect estimates are largely consistent when considering pleiotropy-robust MR implementations like Weighted Median^31^ and MR-Egger^32^, though both come at a cost of statistical power (**Supplementary Methods, Supplementary Table 5-9**).

### Skip-structure accounts for PRS Pleiotropy difference between WorstEpisode and PHQ9 symptoms

As MR results show that PHQ9 symptoms are more likely endorsed due to non-MDD phenotypes that index general dysphoria than MDD, we further ask if this means that genetic studies on PHQ9 symptoms are less likely to lead us to identify MDD-specific biology. To do this we computed 10-fold cross-validated PRS on all PHQ9 and WorstEpisode symptoms in UKBiobank, and used these to predict MDD and non-MDD phenotypes explored above, obtaining the ratio between their prediction R^2^ (PRS Pleiotropy = R^2^_non-MDD_/R^2^_MDD_)^33^. A higher PRS Pleiotropy means lower specificity of a PRS for MDD. For the MDD phenotype, we use LifetimeMDD^30^ in UKBiobank (**Figure 5, Supplementary Table 10**) as well as ICD10-based MDD in iPSYCH2012^28^ and iPSYCH2015i^29^ (**Supplementary Figures 5**,**6, Supplementary Tables 11**,**12**).

**Figure 5.**
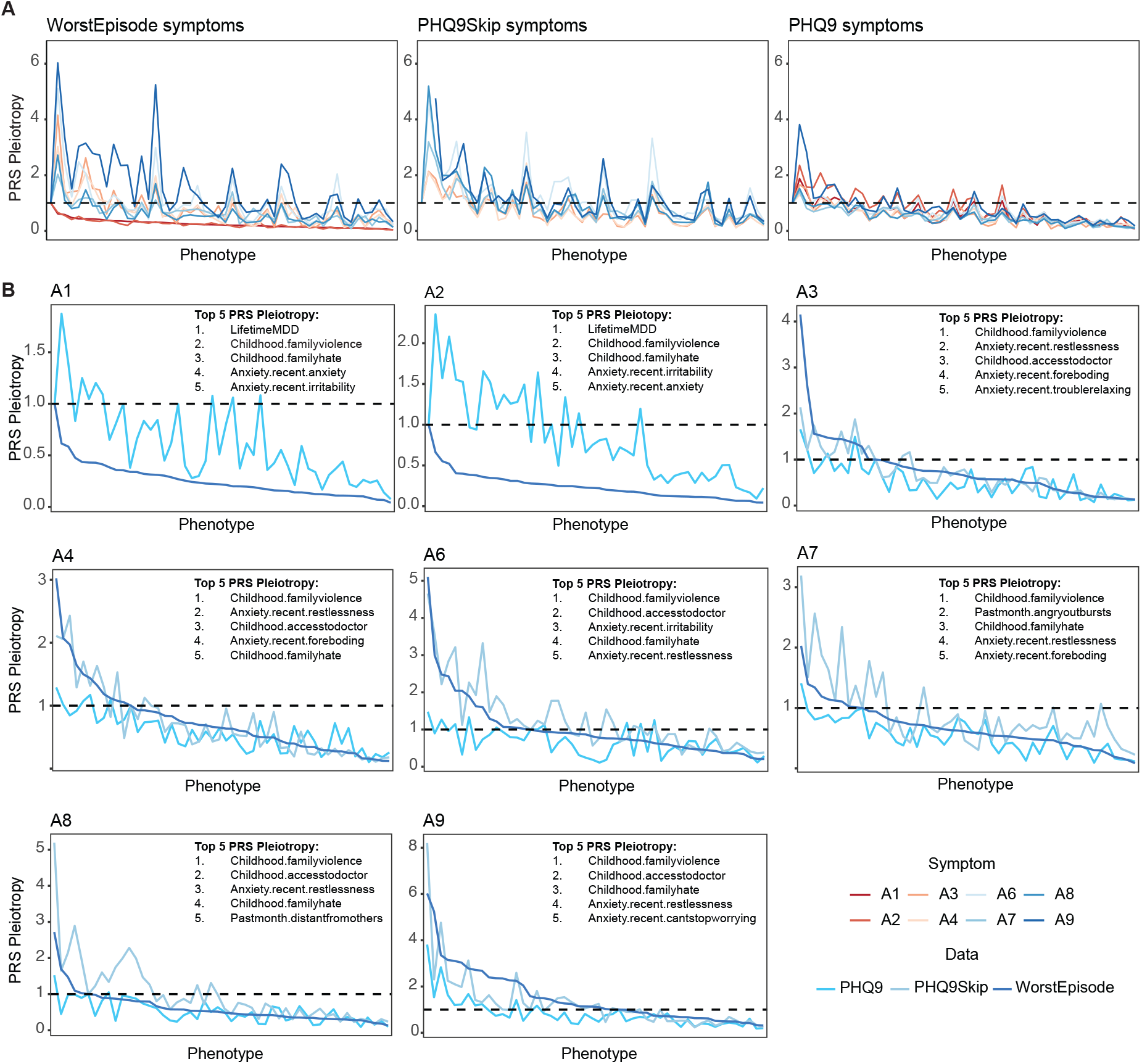
PRS Pleiotropy of PHQ9 and WorstEpisode symptoms. **A**. Mean PRS Pleiotropy for MDD across 10-fold cross-validation of WorstEpisode, PHQ9 and PHQ9Skip symptoms on 50 phenotypes (including LifetimeMDD and 50 non-MDD phenotypes, PRS Pleiotropy = R^2^_non-MDD_/R^2^_MDD_); the MDD phenotype here is LifetimeMDD^30^ defined in UKBiobank; phenotypes on the x-axis across all three panels are ordered by WorstEpisode A1 PRS Pleiotropy in descending order. **B**. Mean PRS Pleiotropy for each symptom across 10-fold cross validation; phenotypes on the x-axis in each panel are ordered by the WorstEpisode symptom PRS Pleiotropy in descending order; top 5 phenotypes in terms of PRS Pleiotropy are indicated.

Overall, we find that WorstEpisode cardinal symptoms A1 (sad mood) and A2 (anhedonia) show lower PRS Pleiotropy across all examined non-MDD phenotypes than their PHQ9 counterpart (**Figure 5A**,**B, Supplementary Figures 5**,**6**), but the non-cardinal WorstEpisode (A3-A9) symptoms show much higher PRS Pleiotropy than the corresponding PHQ9 symptoms (**Figure 5A**,**B, Supplementary Figures 5**,**6**). This is in contrast to most of our results from the MR analyses, where we show that PHQ9 symptoms are more highly associated with genetic effects on non-MDD phenotypes. We hypothesise that this difference must come from how either sets of symptoms predict MDD, which requires the endorsement of either symptoms A1 or A2: WorstEpisode symptoms A3-A9, assessed with a skip-structure conditional on endorsement of symptoms A1 or A2, cannot predict the genetic liability in MDD shared with symptoms A1 and A2. In contrast, PHQ9 symptoms A3-A9 are not assessed with a skip-structure, and therefore may be able to predict those components of MDD genetic liability that are shared with PHQ9 symptoms A1 and A2. This would be in agreement with our MR finding that PHQ9 symptoms are more associated with genetic effects on MDD.

To verify this, we first obtain rG between MDD (LifetimeMDD^30^, and MDD assessed in PGC29^27^, iPSYCH2012^28^ and iPSYCH2015i^29^, **Supplementary Methods**) and both sets of symptoms. Consistent with our hypothesis, WorstEpisodes A1 and A2 have higher rG with MDD across all four cohorts than the corresponding PHQ9 symptoms, while it is generally the reverse for symptoms A3-A9 (**Supplementary Figure 7**). We then ask if this difference is reduced between WorstEpisode symptoms A3-A9 and PHQ9Skip symptoms A3-A9 (PHQ9 symptoms with skip-structure applied), and find that this is generally true, though error bars are large for some PHQ9Skip symptoms (**Supplementary Figure 7**). Third, we perform factor analyses on genetic covariance between MDD and both sets of symptoms to identify their genetic sharing using genomicSEM^8^. We find that a two-factor model best captures sharing between WorstEpisode symptoms and MDD, where the first factor loads onto MDD, A1 and A2, and the second loads onto A3 to A9. In contrast, a one-factor model captures genetic sharing between PHQ9 symptoms and MDD best. Once a skip-structure is applied, the wide-spread sharing between MDD and all PHQ9 symptoms is gone in all tested cohorts (**Supplementary Figure 8**). Finally, we obtain PRS Pleiotropy of PHQ9Skip symptoms A3-A9 and find that once the skip-structure is applied, PHQ9Skip symptoms A3-A9 have much higher PRS Pleiotropy for all non-MDD phenotypes across the board (**Figure 5A**,**B, Supplementary Figures 5**,**6**). This is consistent with our hypothesis. In other words, because of the skip-structure, WorstEpisode symptoms A3-A9 do not capture a large proportion of genetic liability to MDD.

In fact, WorstEpisode symptoms A3-A9 capture genetic components indexing different liabilities within individuals enriched for MDD (through their endorsement of A1 and A2). WorstEpisode symptoms A3-A9 show the highest PRS Pleiotropy for childhood trauma, including violence in the family and having no access to a doctor, as well as items in the GAD7 questionnaire (**Figure 5B**), with some differences in magnitude between individual items. This demonstrates that WorstEpisode symptoms capture potential genetic heterogeneity among individuals with cardinal symptoms of MDD in relation to other non-MDD conditions. While PHQ9 symptoms A3-A9 show similar PRS Pleiotropy trends as WorstEpisodes A3-A9 (**Figure 5B**), they capture this heterogeneity to a much smaller extent.

### WorstEpisode symptoms are better at capturing within-MDD heterogeneity

Finally, we investigate the utility of WorstEpisode and PHQ9 symptoms to identify genetically driven symptom dimensions of MDD with genomicSEM^8^. We find that rG (and genetic covariance, **Supplementary Methods**) among PHQ9 symptoms are significantly higher (mean rG = 0.80, sd = 0.14, **Figure 6A**) than those among WorstEpisode symptoms (mean rG = 0.40, sd = 0.27, **Figure 6A**) indicating that the WorstEpisode symptoms, especially A3-A9, are more genetically heterogeneous than PHQ9 symptoms. The choice between using either set of symptoms in factor analysis for identifying underlying symptom dimensions of MDD subtypes is therefore likely to make an important difference.

**Figure 6.**
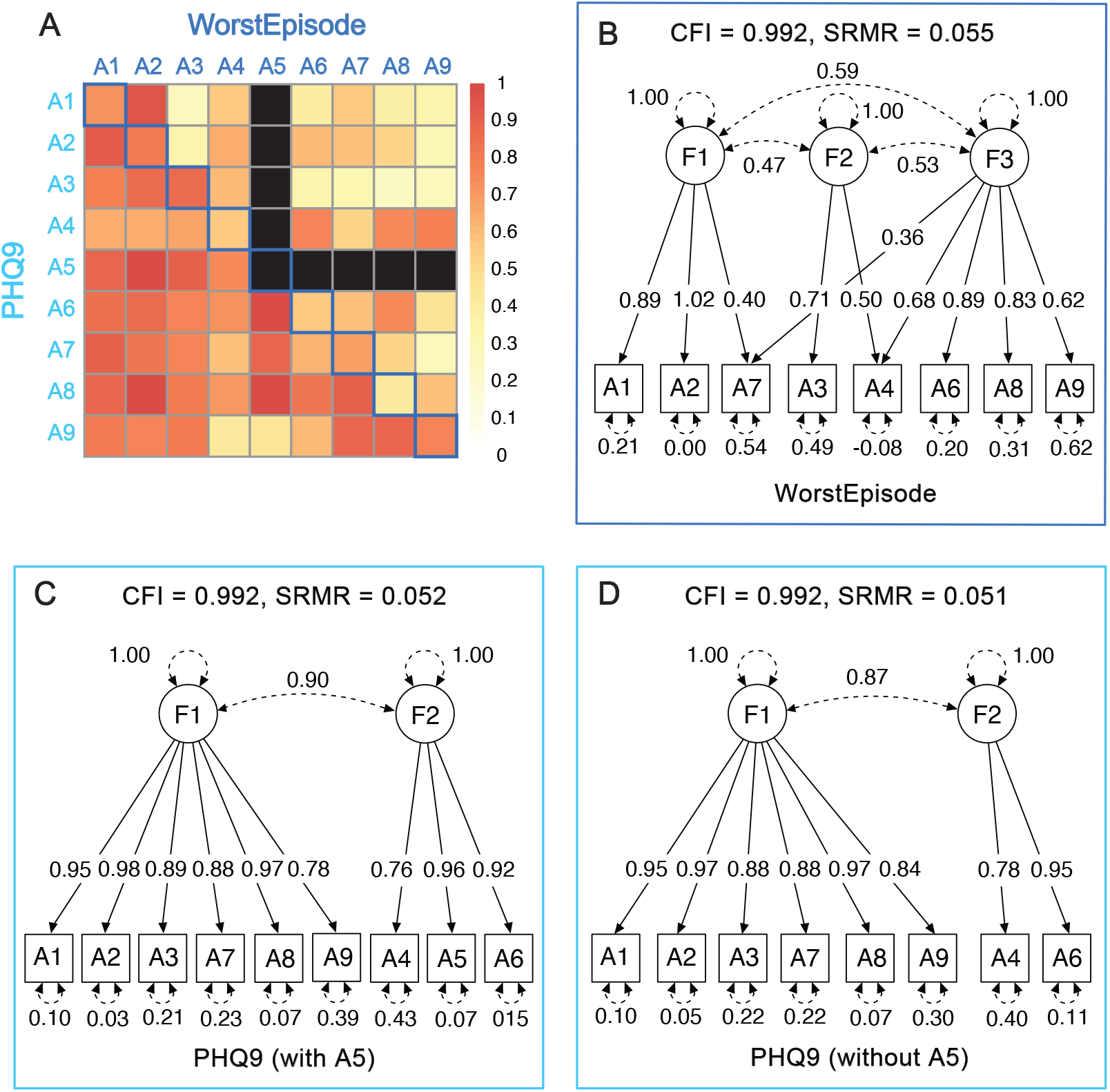
rG estimates and factor analysis of PHQ9 and WorstEpisode symptoms of MDD. **A**. rG estimates between pairs of symptoms. Blue squares on the diagonal line show rG estimates between pairs of corresponding WorstEpisode and PHQ9 symptoms, squares in the upper triangle show rG estimates between pairs of WorstEpisode symptoms, squares in the lower triangle show rG estimates between pairs of PHQ9 symptoms. Black squares indicate no data for WorstEpisode symptom A5. **B**. CFA factors loadings on WorstEpisode symptoms. **C**. CFA factor loadings on PHQ9 symptoms, including symptom A5. **D**. CFA factor loadings on PHQ9 symptoms, excluding symptom A5. Only models with P < 0.05 are considered in model comparisons; models with best fit are shown.

We first perform an exploratory factor analysis (EFA, **Methods**) on the genetic covariance matrix of the 8 WorstEpisode symptoms (missing A5), obtaining results for solutions based on one to three factors. We then run follow-up confirmatory factor analyses (CFA, **Methods**) on the EFA solutions (standardised loadings > 0.2 are retained). The two-factor solution (χ2=57.36; AIC=91.37; CFI=0.987; SRMR=0.078) and three-factor solution (χ2=39.74; AIC=81.74; CFI=0.991; SRMR=0.055) both give much better fits than the single, common factor model (χ2=249.87; AIC=277.87; CFI=0.915; SRMR=0.137). We choose the three-factor solution for its fit and the low correlations between its factors (range 0.47 - 0.59, **Figure 6B**). In this solution, the first factor loads onto A2, A1 and A7 (all symptoms are ordered by loadings), the second loads onto A3 and A4, while the third loads onto A6, A8, A4, A9 and A7. This is consistent with previous factor analyses based on phenotypic covariances which, though not always consistent with each other^34^, generally identify factors that group dysphoric, neurovegetative, and psychomotor symptoms/signs of MDD^12,35,36^.

We then perform the same analyses on the genetic covariance matrix of the PHQ9 symptoms (**Figure 6C**). The two factor solution (χ2=44.34; AIC=82.34; CFI=0.993; SRMR=0.052) only marginally outperform the one-factor solution (χ2=54.09; AIC=90.09; CFI=0.990; SRMR=0.062), and the three factor model did not yield any standardised loadings greater than 0.2. Removing A5 from PHQ9, to make this analysis comparable with that in the WorstEpisode symptoms, yields similar results (**Supplementary Methods**). In the two-factor solutions, the first factor loads onto A2, A8, A1, A3, A7, and A9, and the second factor loads onto A5 (where it is in the model), A6, and A4 (**Figure 6D**). This structure is inconsistent with that derived from the WorstEpisode symptoms; notably, the correlation between factors 1 and 2 is much higher (0.90) than the average correlation between factors observed in WorstEpisode symptoms (0.53), reflective of the high correlations between all PHQ9 symptoms. In other words, PHQ9 symptoms do not capture as much heterogeneity as WorstEpisode symptoms, and are not interchangeable in their utility for identifying genetically driven symptom dimensions of MDD.

## Discussion

In this paper, we examine whether symptom-level data assessed in the general population by the PHQ9 capture the same biology as WorstEpisode MDD symptoms assessed through the CIDI-SF. We find that while they have similar h^2^_SNP_, they have distinct genetic components, and this difference can only partly be accounted for by the skip-structure of the CIDI-SF or the severity threshold for symptom endorsement in PHQ9. We further find that PHQ9 symptoms are much more genetically correlated with each other than WorstEpisode symptoms, and factor analysis on their genetic covariance matrices do not identify the same underlying symptom dimensions for MDD. The two sets of symptoms are not interchangeable in genetic analyses; they lead to different findings with different biological meanings.

Some of the differences between the two sets of symptoms are due to the implementation of the skip-structure in CIDI-SF when assessing WorstEpisode symptoms. We find that non-cardinal WorstEpisode symptoms, which are only assessed when cardinal symptoms A1 and A2 are endorsed, are only able to capture those genetic components of MDD not shared with A1 and A2. As such, their PRS Pleiotropy on 50 non-MDD phenotypes are higher than non-cardinal PHQ9 symptoms, which are not subject to the skip-structure. Once the skip-structure is applied in non-cardinal PHQ9 symptoms, most of their differences from the corresponding WorstEpisode symptoms are gone. The remaining differences may be due to recall differences between current symptoms and those during a potentially distant MDD episode. Overall, we find that the skip-structure effectively reduces the genetic sharing between non-cardinal WorstEpisode symptoms and MDD, while increasing their ability to capture heterogeneity amongst those with MDD. Of particular interest, the non-cardinal WorstEpisode symptoms show highest PRS Pleiotropy for childhood trauma and anxiety symptoms, pointing to them as potential axes of genetic heterogeneity among those likely with MDD.

WorstEpisode symptoms must, by definition, be occurring during MDD episodes. Most PHQ9 will not be, and this can explain most of our results: PHQ9 symptoms show more genetic sharing with more stable traits like neuroticism, insomnia and measures of subjective well-being. In other words, PHQ9 symptoms index general dysphoria more than episodic MDD. As pointed out in “The Clinician’s Illusion” ^37^, if one samples at any given time the population currently suffering from a condition (a point prevalence sample), one is more likely to oversample those who have long episodes of illness. Our findings on PHQ9 symptoms are consistent with this hypothesis: they index traits like chronic insomnia more than the episodic state of MDD, a recurrent and relapsing disorder. Our findings also complement previous findings that self-ratings with PHQ9 likely lead to the inclusion of long-standing conditions as well as those due to external causes unrelated to MDD, and this can inflate MDD prevalence^15,16^. This may be exacerbated by the “healthy volunteer” effect in the general population that answers the MHQ in the UKBiobank^2,38^. This does not discredit the PHQ9 as a sensitive screening instrument for current MDD, especially in ruling out those without MDD, or as a measurement of depression severity, as it is intended for^3^. It may not, however, be suitable for identifying symptom dimensions in MDD patients. As previously argued, “not all instruments are appropriate for all purposes” ^39,40^.

Our results should be interpreted in the context of the following limitations. First, our sample sizes are low, leading to low statistical power in GWAS and MR analyses. This can be improved with increased data collection at the symptom level, using methods going beyond diagnostic questionnaires^40,41^. Second, genetic associations identified for PHQ9 or WorstEpisode symptoms may be due to collider bias from the ascertainment of individuals participating in the MHQ: participation in the MHQ has positive rGs with higher educational attainment and better health, and negative rGs with psychological distress and schizophrenia^38^. Third, no clinician ratings are available in UKBiobank to be compared like-to-like with PHQ9 and CIDI-SF ratings, and hence we do not have insights into biases inherent in self-rated symptoms. The latter, in particular, may suffer from greater recall-bias. Both these limitations may be improved in truly representative population-based, clinician-assessed cohorts such as electronic health records or national registries, especially when clinicians’ notes are available to assess symptom-level disease characteristics. Finally, factors identified through genomicSEM should not necessarily be seen as “real” or “entity-like” ^10^, as they may be subject to statistical underdetermination^42,43^. Rather, they reflect genetic sharing among PHQ9 and WorstEpisode symptoms, and greater utility of WorstEpisode symptoms to detect symptom dimensions of episodic MDD is determined only through similarities between its clustering and previous results from phenotypic factor analyses.

In summary, our work explicitly examines two instruments (PHQ9 and CIDI-SF) widely used in disease cohorts and biobanks to assess MDD symptoms, and which have been previously used to understand MDD heterogeneity^7,9^. We find that symptoms assessed through these instruments capture different underlying biology, and are not interchangeable in genetic analysis. In particular, PHQ9 symptoms index general dysphoria more than episodic MDD, and WorstEpisode symptoms are more suitable for investigations into symptom dimensions of MDD.

## Methods

### Definition of WorstEpisode and PHQ9 MDD symptoms in UKBiobank

Individual-level MDD symptom data are available for UKBiobank participants from those who answered the questions for MDD symptoms in the CIDI-SF and PHQ9 conducted through an online mental health follow-up survey (MHQ, data category 138). Details of definitions of each WorstEpisode and PHQ9 symptoms can be found in **Supplementary Methods**, and sample sizes can be found in **Supplementary Tables 1** and **2** respectively. Definitions and sample sizes of PHQ9Strict and PHQ9Skip symptoms are described in **Supplementary Methods** and detailed in **Supplementary Table 2**. To investigate PHQ9 and WorstEpisode symptoms at the same GWAS power, we perform downsampling: for each corresponding pair of PHQ9 and WorstEpisode symptoms, we downsample the one with higher effective sample sizes accounting for imbalance between cases and controls (N_eff_ = 4/(1/N_cases_ + 1/N_controls_) to the same N_eff_ of the one with lower N_eff_ (**Supplementary Table 1, 2**), keeping its prevalence unchanged.

### Definition of other phenotypes in UKBiobank

We further selected other phenotypes, including insomnia, measures of subjective well-being, neuroticism, individual neuroticism items, anxiety symptoms and stressful life event exposures in the UKBiobank to test for genetic sharing with PHQ9 and WorstEpisode symptoms. For all data fields in UKBiobank, endorsement criteria and sample sizes, see **Supplementary Table 4**.

### Genome-wide associations in UK Biobank

Genome-wide association analysis was performed using imputed genotype data at 5,776,313 SNPs (MAF >= 0.05, INFO score >= 0.9) in PLINK2^44^. We used 20 PCs computed with flashPCA^45^ on 337,198 White-British individuals in UK Biobank and genotyping arrays as covariates, using a logistic regression model for binary phenotypes and a linear regression model for continuous phenotypes.

### GWAS on MDD from PGC29 and iPSYCH cohorts

We used the publicly available PGC29 GWAS summary statistics from Psychiatric Genomics Consortium^27^ (https://www.med.unc.edu/pgc/download-results). For iPSYCH, we performed GWAS using logistic regression in PLINK2^44^ on MDD defined by at least one specialty psychiatric care contact registered in the Danish Psychiatric Central Research Register (PCR)^46^ or the Danish National Patient Register (DNPR)^47^ for ICD10 code of F32 or F33, in two independent iPSYCH cohorts iPSYCH2012^28^ and iPSYCH2015i^29^: with 42,250 and 23,351 unrelated individuals with European genetic ancestry respectively. We used 5,210,642 and 5,222,714 SNPs (MAF >= 0.05, Beagle DR2 >= 0.9, P-value for HWE violation > 10-6) for GWAS in iPSYCH2012 and iPSYCH2015i respectively. We used the top 10 genomic PCs from individuals in iPSYCH2012 and iPSYCH2015i, computed using FlashPCA^45^ as covariates to control for population structure in each of the cohorts. Details of the iPSYCH cohorts can be found in **Supplementary Methods**.

### SNP-heritability and genetic correlation

To test for heritability of each symptom and the rG between pairs of symptoms, we performed LD score regression implemented in LDSC v1.0.1^13^, using in-sample LD scores estimated from 10,000 random White British UKBiobank^1^ individuals at SNPs with MAF > 0.05 as reference. We assumed the population prevalence of each symptom was equal to its sample prevalence in UKBiobank, then estimated SNP-heritability on the liability scale for each symptom and rG between pairs of symptoms. One-sided paired t-tests are conducted on rGs between the two sets of symptoms and non-MDD phenotypes in R.

### Univariable Mendelian Randomization (UVMR)

Two-sample UVMR was performed using MendelianRandomization v0.6.0^25^ implemented in R v4.0.3. For each pair of exposures and outcomes, we used SNPs that were genome-wide significantly (P-value < 5×10^−6^) associated with each exposure as instruments. Clumping and LD pruning were performed with default settings with R library *ieugwasr*: clump_kb=10,000, clump_r2=0.001. We tested the validity of instruments used in MR using the F statistics (**Supplementary Tables 5**,**6**,**8**). Multiple-testing corrections on the number of symptoms were performed separately for PHQ9 and WorstEpisode symptoms. To assess horizontal pleiotropy^48^, we further conducted pleiotropy-robust Weighted Median MR^31^ and MR Egger^32^ to compare the MR estimates between different MR models (**Supplementary Methods**). One-sided paired t-tests are conducted on effect sizes from UVMR analyses in R.

### Multivariable MR Bayesian model averaging (MR-BMA)

We use MR-BMA^26^, a statistical learning algorithm for two-sample multivariable MR in order to select likely causal exposures from a larger set of candidate exposures. We selected independent genetic variants associated with any of the symptoms as instrumental variables for the multivariable MR model. We assumed that half of the tested items were expected causal risk factors (prior = 0.5) when iterating through all possible combinations of candidate models in the model averaging algorithm. We ranked symptoms according to their marginal inclusion probability and calculated the respective empirical P-values. Finally, we adjusted for multiple testing using the Benjamini Hochberg false discovery rates (FDR). Only those exposures with an FDR corrected P-value below 0.05 were significant and therefore being reported in this paper.

### PRS prediction and PRS Pleiotropy

For all in-sample PRS prediction in UKBiobank, we perform 10-fold cross-validation PRS on all PHQ9 and WorstEpisode symptoms in UKBiobank, by performing GWAS on each symptom 10 times, each time using 90% of the individuals, and building PRS from this GWAS results with PRSice v2^49^. We evaluated predictive accuracy for observed LifetimeMDD and 50 non-MDD phenotypes that index neuroticism, anxiety, and recent and lifetime stress (**Supplementary Table 4**) in the held-out 10% for all 10 folds. For all PRS predictions in UKBiobank phenotypes, we used 20 genomic PCs and the genotyping array used as covariates. For out-of-sample predictions of MDD diagnostic code in iPSYCH2012 and 2015i, we use PRS built from the same 10-fold GWAS on symptoms in UKBiobank. We perform the prediction on MDD defined by ICD10 code of F32 or F33, in two independent iPSYCH cohorts iPSYCH2012^28^ and iPSYCH2015i^29^, as described in **Supplementary Methods**. We use top 10 genomic PCs from individuals in iPSYCH2012 and iPSYCH2015i as covariates to control for population structure in each of the cohorts in PRS predictions. For binary phenotypes, including LifetimeMDD in UKBiobank and MDD in iPSYCH, we evaluated accuracy using Nagelkerke’s *R*^2^. For all quantitative phenotypes, including neuroticism, we evaluated accuracy using ordinary *R*^2^. PRS Pleiotropy^33^ is calculated for each PHQ9 and WorstEpisode symptom using the ratio of its PRS predictions on 50 non-MDD phenotypes and its prediction on LifetimeMDD in UKBiobank or ICD10-based MDD in iPSYCH cohorts (PRSPleiotropy = *R*^2^_non-MDD_/*R*^2^_MDD_, **Supplementary Tables 10-12**).

### Factor analysis on genetic covariances using genomicSEM

The genetic exploratory factor analysis (EFA) was conducted using the *psych* library in R with the minimum residual (minres) extraction approach and “promax” rotation enabled by the GPArotation, on PHQ9 symptoms and WorstEpisode symptoms respectively using genetic covariance matrices estimated with LDSC. Solutions with one to three factors were carried forward and assessed for their model fit (retaining factor loadings >0.2) using confirmatory factor analysis (CFA) in genomicSEM^8^; only models with significant fit (p<0.05) are considered in model comparisons.

## Author contributions

LH and NC wrote the paper. VZ, KK and NC designed the study. ST supported the MR analyses. MK, VA, TW, and AJS supported iPSYCH analyses. LH and NC performed all analyses. All authors reviewed the paper.

## Data Availability

UK Biobank genotype and phenotype data used in this study are from the full release of the UKBiobank Resource obtained under application no. 28709. We used publicly available summary statistics from PGC29 (https://www.med.unc.edu/pgc/results-and-downloads). The individual-level data from the iPSYCH cohort is not publicly available due to institutional restrictions on data sharing and privacy concerns. Summary statistics of all PHQ9 and WorstEpisode symptoms presented in this paper are available on https://doi.org/10.6084/m9.figshare.22041212.

## Ethical approval

This research was conducted under ethical approval from the UK Biobank Resource under application no. 28709. The use of iPSYCH data follows the standards of the Danish Scientific Ethics Committee, the Danish Health Data Authority, the Danish Data Protection Agency, and the Danish Neonatal Screening Biobank Steering Committee. Data access was via secure portals in accordance with Danish data protection guidelines set by the Danish Data Protection Agency, the Danish Health Data Authority, and Statistics Denmark.

## Code availability

Publicly available tools that are used in data analyses are described wherever relevant in Methods and Reporting Summary. Custom code for softImpute imputation of the MDD-relevant phenome and calculating PRS Pleiotropy are available at https://github.com/caina89/MDDSymptoms.

## Acknowledgements

LH, ST, VZ and NC are supported by the TUM Global Incentive Fund. The iPSYCH team is supported by grants from the Lundbeck Foundation (R102-A9118, R155-2014-1724, and R248-2017-2003), NIMH (1R01MH124851-01) and the Universities and University Hospitals of Aarhus and Copenhagen. The Danish National Biobank resource is supported by the Novo Nordisk Foundation. High-performance computer capacity for handling and statistical analysis of iPSYCH data on the GenomeDK HPC facility is provided by the Center for Genomics and Personalised Medicine and the Centre for Integrative Sequencing, iSEQ, Aarhus University, Denmark. AJS is supported by Lundbeckfonden Fellowship R335-2019-2318. VA is supported by Lundbeck foundation postdoctoral grant R380-2021-1465. ST is supported by a Medical Research Council UK PhD Studentship. The authors thank Jonathan Flint, Noah Zaitlen, Joel Mefford, Andrew Dahl, Richard Border and Eiko Fried for constructive discussions and feedback on the paper.

## References

1. Bycroft, C. et al. The UK Biobank resource with deep phenotyping and genomic data. Nature 562, 203–209 (2018).

2. Davis, K. A. S. et al. Mental health in UK Biobank - development, implementation and results from an online questionnaire completed by 157 366 participants: a reanalysis. BJPsych Open 6, e18 (2020).

3. Kroenke, K., Spitzer, R. L. & Williams, J. B. The PHQ-9: validity of a brief depression severity measure. J. Gen. Intern. Med. 16, 606–613 (2001).

4. Diagnostic and Statistical Manual of Mental Disorders: Dsm-5. (Amer Psychiatric Pub Incorporated, 2013).

5. Kessler, R. C., Andrews, G., Mroczek, D., Ustun, B. & Wittchen, H.-U. The World Health Organization Composite International Diagnostic Interview short-form (CIDI-SF). International Journal of Methods in Psychiatric Research vol. 7 171–185 Preprint at https://doi.org/10.1002/mpr.47 (1998).

6. Levinson, D. et al. Brief Assessment Of Major Depression For Genetic Studies: Validation Of Cidi-Sf Screening With Scid Interviews. European Neuropsychopharmacology vol. 27 S448 Preprint at https://doi.org/10.1016/j.euroneuro.2016.09.514 (2017).

7. Thorp, J. G. et al. Genetic heterogeneity in self-reported depressive symptoms identified through genetic analyses of the PHQ-9. Psychol. Med. 50, 2385–2396 (2020).

8. Grotzinger, A. D. et al. Genomic structural equation modelling provides insights into the multivariate genetic architecture of complex traits. Nat Hum Behav 3, 513–525 (2019).

9. Thorp, J. G. et al. Symptom-level modelling unravels the shared genetic architecture of anxiety and depression. Nat Hum Behav 5, 1432–1442 (2021).

10. van Loo, H. M., Aggen, S. H. & Kendler, K. S. The structure of the symptoms of major depression: Factor analysis of a lifetime worst episode of depressive symptoms in a large general population sample. J. Affect. Disord. 307, 115–124 (2022).

11. Li, Y. et al. The structure of the symptoms of major depression: exploratory and confirmatory factor analysis in depressed Han Chinese women. Psychol. Med. 44, 1391–1401 (2014).

12. Kendler, K. S., Aggen, S. H. & Neale, M. C. Evidence for multiple genetic factors underlying DSM-IV criteria for major depression. JAMA Psychiatry 70, 599–607 (2013).

13. Bulik-Sullivan, B. K. et al. LD Score regression distinguishes confounding from polygenicity in genome-wide association studies. Nat. Genet. 47, 291–295 (2015).

14. Bulik-Sullivan, B. et al. An atlas of genetic correlations across human diseases and traits. Nat. Genet. 47, 1236–1241 (2015).

15. von Glischinski, M., von Brachel, R., Thiele, C. & Hirschfeld, G. Not sad enough for a depression trial? A systematic review of depression measures and cut points in clinical trial registrations. J. Affect. Disord. 292, 36–44 (2021).

16. Levis, B. et al. Patient Health Questionnaire-9 scores do not accurately estimate depression prevalence: individual participant data meta-analysis. J. Clin. Epidemiol. 122, 115–128.e1 (2020).

17. Havinga, P. J. et al. Doomed for Disorder? High Incidence of Mood and Anxiety Disorders in Offspring of Depressed and Anxious Patients: A Prospective Cohort Study. J. Clin. Psychiatry 78, e8–e17 (2017).

18. Kendler, K. S., Gardner, C. O., Gatz, M. & Pedersen, N. L. The sources of co-morbidity between major depression and generalized anxiety disorder in a Swedish national twin sample. Psychol. Med. 37, 453–462 (2007).

19. Kendler, K. S., Karkowski, L. M. & Prescott, C. A. Causal relationship between stressful life events and the onset of major depression. Am. J. Psychiatry 156, 837–841 (1999).

20. Kendler, K. S. & Karkowski-Shuman, L. Stressful life events and genetic liability to major depression: genetic control of exposure to the environment? Psychological Medicine vol. 27 539–547 Preprint at https://doi.org/10.1017/s0033291797004716 (1997).

21. Peterson, R. E. et al. Molecular Genetic Analysis Subdivided by Adversity Exposure Suggests Etiologic Heterogeneity in Major Depression. Am. J. Psychiatry 175, 545–554 (2018).

22. Eysenck, S. B. G., Eysenck, H. J. & Barrett, P. A revised version of the psychoticism scale. Pers. Individ. Dif. 6, 21–29 (1985).

23. Burgess, S. et al. Guidelines for performing Mendelian randomization investigations. Wellcome Open Res 4, 186 (2019).

24. Smith, G. D. & Ebrahim, S. ‘Mendelian randomization’: can genetic epidemiology contribute to understanding environmental determinants of disease? Int. J. Epidemiol. 32, 1–22 (2003).

25. Yavorska, O. O. & Burgess, S. MendelianRandomization: an R package for performing Mendelian randomization analyses using summarized data. Int. J. Epidemiol. 46, 1734–1739 (2017).

26. Zuber, V., Colijn, J. M., Klaver, C. & Burgess, S. Selecting likely causal risk factors from high-throughput experiments using multivariable Mendelian randomization. Nat. Commun. 11, 29 (2020).

27. Wray, N. R. et al. Genome-wide association analyses identify 44 risk variants and refine the genetic architecture of major depression. Nat. Genet. 50, 668–681 (2018).

28. Pedersen, C. B. et al. The iPSYCH2012 case-cohort sample: new directions for unravelling genetic and environmental architectures of severe mental disorders. Mol. Psychiatry 23, 6–14 (2018).

29. Bybjerg-Grauholm, J. et al. The iPSYCH2015 Case-Cohort sample: updated directions for unravelling genetic and environmental architectures of severe mental disorders. bioRxiv (2020) doi:10.1101/2020.11.30.20237768.

30. Cai, N. et al. Minimal phenotyping yields genome-wide association signals of low specificity for major depression. Nat. Genet. 52, 437–447 (2020).

31. Bowden, J., Davey Smith, G., Haycock, P. C. & Burgess, S. Consistent Estimation in Mendelian Randomization with Some Invalid Instruments Using a Weighted Median Estimator. Genet. Epidemiol. 40, 304–314 (2016).

32. Bowden, J., Davey Smith, G. & Burgess, S. Mendelian randomization with invalid instruments: effect estimation and bias detection through Egger regression. Int. J. Epidemiol. 44, 512–525 (2015).

33. Dahl, A. et al. Phenotype integration improves power and preserves specificity in biobank-based genetic studies of MDD. bioRxiv (2022) doi:10.1101/2022.08.15.503980.

34. van Loo, H. M., de Jonge, P., Romeijn, J.-W., Kessler, R. C. & Schoevers, R. A. Data-driven subtypes of major depressive disorder: a systematic review. BMC Med. 10, 156 (2012).

35. Shafer, A. B. Meta-analysis of the factor structures of four depression questionnaires: Beck, CES-D, Hamilton, and Zung. J. Clin. Psychol. 62, 123–146 (2006).

36. Romera, I., Delgado-Cohen, H., Perez, T., Caballero, L. & Gilaberte, I. Factor analysis of the Zung self-rating depression scale in a large sample of patients with major depressive disorder in primary care. BMC Psychiatry 8, 4 (2008).

37. Cohen, P. & Cohen, J. The clinician’s illusion. Arch. Gen. Psychiatry 41, 1178–1182 (1984).

38. Adams, M. J. et al. Factors associated with sharing e-mail information and mental health survey participation in large population cohorts. Int. J. Epidemiol. 49, 410–421 (2020).

39. Patalay, P. & Fried, E. I. Editorial Perspective: Prescribing measures: unintended negative consequences of mandating standardized mental health measurement. J. Child Psychol. Psychiatry 62, 1032–1036 (2021).

40. Fried, E. I., Flake, J. K. & Robinaugh, D. J. Revisiting the theoretical and methodological foundations of depression measurement. Nature Reviews Psychology vol. 1 358–368 Preprint at https://doi.org/10.1038/s44159-022-00050-2 (2022).

41. Freimer, N. B. & Mohr, D. C. Integrating behavioural health tracking in human genetics research. Nat. Rev. Genet. 20, 129–130 (2019).

42. Johnson, K. Realism and uncertainty of unobservable common causes in factor analysis. Nous 50, 329–355 (2016).

43. Romeijn, J.-W. & Williamson, J. Intervention and Identifiability in Latent Variable Modelling. Minds Mach. 28, 243–264 (2018).

44. Chang, C. C. et al. Second-generation PLINK: rising to the challenge of larger and richer datasets. GigaScience vol. 4 Preprint at https://doi.org/10.1186/s13742-015-0047-8 (2015).

45. Abraham, G., Qiu, Y. & Inouye, M. FlashPCA2: principal component analysis of Biobank-scale genotype datasets. Bioinformatics 33, 2776–2778 (2017).

46. Mors, O., Perto, G. P. & Mortensen, P. B. The Danish Psychiatric Central Research Register. Scand. J. Public Health 39, 54–57 (2011).

47. Lynge, E., Sandegaard, J. L. & Rebolj, M. The Danish National Patient Register. Scand. J. Public Health 39, 30–33 (2011).

48. Burgess, S., Foley, C. N. & Zuber, V. Inferring Causal Relationships Between Risk Factors and Outcomes from Genome-Wide Association Study Data. Annu. Rev. Genomics Hum. Genet. 19, 303–327 (2018).

49. Choi, S. W. & O’Reilly, P. PRSice 2: POLYGENIC RISK SCORE SOFTWARE (UPDATED) AND ITS APPLICATION TO CROSS-TRAIT ANALYSES. European Neuropsychopharmacology vol. 29 S832 Preprint at https://doi.org/10.1016/j.euroneuro.2017.08.092 (2019).

