## SupplementaryMaterials for "Polygenic analyses show important differences between MDD symptoms collected using PHQ9 and CIDI-SF"

**Table of Contents**

***Supplementary Methods*** ***3***

Sample filtering in UK Biobank 3

Genotype quality control 3

Control for population structure with principal component analysis 3

Definition of WorstEpisode and PHQ9 symptoms 4

iPSYCH cohort 4

Mendelian Randomization analysis considerations 5

Factor analyses using GenomicSEM 6

***Supplementary Figures*** ***7***

Supplementary Figure 1 7

Supplementary Figure 2 8

Supplementary Figure 3 9

Supplementary Figure 4 10

Supplementary Figure 5 12

Supplementary Figure 6 13

Supplementary Figure 7 14

Supplementary Figure 8 15

***Supplementary Table Legends*** ***16***

Supplementary Table 1. 16

Supplementary Table 2. 16

Supplementary Table 3. 16

Supplementary Table 4. 16

Supplementary Table 5. 16

Supplementary Table 6. 16

Supplementary Table 7. 16

Supplementary Table 8. 17

Supplementary Table 9. 17

Supplementary Table 10-12. 17

***Supplementary References*** ***18***

### Supplementary Methods

#### Sample filtering in UK Biobank

Of all 502,637 samples in UKBiobank[1] full release, we performed the following QC steps to select the samples for use in our analyses. We first removed samples that were not included in the UKBiobank full release PCA analysis, which includes samples that were indicated as "het.missing.outliers" ("Indicates samples identified as outliers in heterozygosity and missing rates, which indicates poor-quality genotypes for these samples"), "excess.relatives" ("Indicates samples which have more than 10 putative third-degree relatives in the kinship table"), and whose "Submitted.Gender" were different from "Inferred.Gender". Applying these filters brought the sample size down to 407,219. We checked that the remaining sample contains only one out of any pair or group of related individuals with relatedness > 0.05. We then selected samples indicated to be "in.white.British.ancestry.subset" ("Indicates samples who self-reported 'White British' and have very similar genetic ancestry based on a principal components analysis of the genotypes"), resulting in a sample size of 337,545. We then removed 337 samples indicated as having "putative.sex.chromosome.aneuploidy" ("Indicates samples identified as putatively carrying sex chromosome configurations that are not either XX or XY"). Finally, we removed 79 samples who have withdrawn their consent for use of their genetic data in analyses, arriving at our final set of 337,129 samples passing QC1. Of these samples, 37,041 were part of UK Biobank Lung Exome Variant Evaluation (UKBiLEVE), a study for chronic obstructive pulmonary disease (COPD). We retain all samples in UKBiLEVE, but as they are genotyped using a custom array optimised for coverage over regions implicated in lung health and disease, we consistently use 'genotyping array' as a covariate in all our analyses.

#### Genotype quality control

We performed stringent filtering on imputed variants (version 3) used for GWAS in this study, removing all insertions and deletions (INDELs) and multi-allelic SNPs: we hard-called genotypes from imputed dosages at 9,720,420 biallelic SNPs with imputation INFO score greater than 0.9, MAF greater than 0.1%, and P value for violation of Hardy-Weinberg equilibrium > $10^{-6}$, in individuals with a genotype probability threshold of 0.9 (individuals with genotype probabilities below 0.9 would be assigned a missing genotype). Of these, 5,776,313 SNPs are common (MAF > 5%). We consistently use these SNPs for all analyses in this study.

#### Control for population structure with principal component analysis

We performed principal component analysis (PCA) on directly genotyped SNPs from samples in UKBiobank and used PCs as covariates in all our analyses to control for population structure using flashPCA[2]. From the array genotype data, we first removed all samples who did not pass QC, leaving 337,129 White-British, unrelated samples. We then removed SNPs not included in the phasing and imputation and retained those with minor allele frequencies (MAF) >= 0.1%, and P value for violation of Hardy-Weinberg equilibrium > $10^{-6}$, leaving 593,300 SNPs. We then removed 20,567 SNPs that are in known structural variants (SVs) and the major histocompatibility complex (MHC) as recommended by UKBiobank[1], leaving 572,733 SNPs. Of these, 334,702 are common (MAF > 5%), and from these common SNPs we further filtered based on missingness <0.02 and pairwise LD r2 < 0.1 with SNPs in a sliding window of 1000 SNPs to obtain 68,619 LD-pruned SNPs for computing PCs using flashPCA.

We obtained 20 PCs, their eigenvalues, loadings and variance explained, and consistently use these PCs as covariates for all our genetic analyses.

**Definition of WorstEpisode and PHQ9 symptoms**

Of the nine symptoms, two consist of independent subgroups. A3 ("Did you gain or lose weight without trying, or did you stay about the same weight?", data field 20536) consists of A3a ("Gained weight"), A3b ("Lost weight") and A3c ("Both gained and lost some weight during the episode"). A4 ("Did your sleep change?", data field 20532) consists of A4a ("Was that: [re sleep change] Trouble falling asleep?", data field 20533), A4b ("Was that: [re sleep change] Sleeping too much?", data field 20534) and A4c ("Was that: [re sleep change] Waking too early?", data field 20535). As such, we have defined a total of 14 WorstEpisode symptoms, including these subgroups, for further analysis (sample size and criteria see **Supplementary table 1**).

For PHQ9 symptoms, we counted participants who indicated having experienced each symptom at any frequency as "cases", and only participants who indicated having never experienced it as "controls" (**Supplementary Table 2**). To test the influence of skip-structure on PHQ9 symptoms, we conditioned 7 out of 9 (A3-A9) questions in the PHQ on the stem questions A1 "Recent feelings of depression" (data field 20510), and A2 "Recent lack of interest or pleasure in doing things" (data field 20514, sample size and criteria see **Supplementary Table 2**). These are defined as PHQ9Skip symptoms. Similarly, to impose a stricter criteria for symptom endorsement following DSM5 on the PHQ9 symptoms, we required a frequency of "nearly everyday" for symptom endorsement (sample size and criteria see **Supplementary Table 2**). These are defined as PHQ9Strict symptoms.

We also performed a skip-structure on PHQ9Strict by conditioning questions A3-A9 on the screening questions A1 and A2, but their sample sizes are small (sample size and criteria see **Supplementary Table 2**). We are therefore unable to obtain $h^2_{SNP}$ and rG for PHQ9Strict symptoms with skip-structure.

**iPSYCH cohort**

We conducted GWAS on two independent cohorts from the Integrative Psychiatric Research Consortium (iPSYCH) cohort (2012 and 2015i).

The Lundbeck Foundation initiative for Integrative Psychiatric Research (iPSYCH)[3,4] is a case-cohort study of all singleton births between 1981 and 2008 to mothers legally residing in Denmark and who were alive and residing in Denmark on their first birthday (N=1,657,449). The iPSYCH 2015 case-cohort comprises two enrollments from this base population. The iPSYCH 2012 case-cohort enrolled 86,189 individuals (30,000 random population controls; 57,377 psychiatric cases)[3]. The iPSYCH 2015i case-cohort expanded enrollment by an additional 56,233 individuals (19,982 random population controls; 36,741 psychiatric cases)[3,4]. DNA was extracted from dried blood spots stored in the Danish Neonatal Screening Biobank[5] and genotyping was performed on the Infinium PsychChip v1.0 array (2012) or the Global Screening Array v2 (2015i). Psychiatric diagnoses were obtained from the Danish Psychiatric Central Research Register (PCR)[6] and the Danish National Patient Register (DNPR)[7]. Diagnoses in these registers are made by licensed psychiatrists during in- or out- patient specialty care but diagnoses or treatments assigned in primary care are not included. Linkage across population registers, to parents where known, and to the neonatal biobank is possible via unique citizen identifiers of the Danish Civil Registration System[8]. The use of this data follows standards of the Danish Scientific Ethics Committee, the Danish Health Data Authority, the Danish Data Protection Agency, and the Danish Neonatal Screening Biobank Steering Committee. Data access was via secure portals in accordance with Danish data protection guidelines set by the Danish Data Protection Agency, the Danish Health Data Authority, and Statistics Denmark. For this study, we use an unrelated, homogeneous ancestry subset of these data from the 2012 and 2015i cohorts. We include individuals with available genotypes, that passed our quality control, and were either a random control or diagnosed with major depressive disorder (MDD) as of Dec 31, 2015 (2012: Ncontrols = 23,371, Ncases = 18,879; 2015i: Ncontrols = 15,163, Ncases = 8,188; Total: Ncontrols = 38,534, Ncases = 27,067).

Genotype phasing, imputation, and quality control were performed in parallel in the 2012 and 2015i cohorts according to custom, mirrored protocols. Briefly, phasing and imputation were conducted using BEAGLEv5.1[9,10], both steps including reference haplotypes from the Haplotype Reference Consortium v1.1 (HRC)[11]. Quality control was applied prior to and following imputation to correct for missing data across SNPs and individuals, SNPs showing deviations from Hardy-Weinberg equilibrium in controls, abnormal heterozygosity of samples, genotype-phenotype sex discordance, minor allele frequency (MAF), batch artifacts, and imputation quality. Kinship was detected within and across 2012 and 2015i cohorts using KING[12], censoring to ensure no second degree or higher relatives remained. Ancestry was examined using the smartpca module of EIGENSOFT[13], and PCA outliers from the set of iPSYCH individuals with both grandparents and four grandparents born in Denmark were excluded.

Using 5,210,642 and 5,222,714 SNPs (MAF >= 0.05, Beagle DR2 >= 0.9, P value for HWE violation > $10^{-6}$) on 42,250 individuals in iPSYCH2012 and 23,351 individuals in iPSYCH 2015i respectively, we calculated PRS for each of the 15 summary statistics (phenotyped, imputed and MTAG GWAS) using PRSice v2[14], using the options --clump-kb 250kb --clump-p 1 --clump-r2 0.1 --interval 5e-05 --lower 5e-08. We used the top 10 genomic PCs from all 42,250 and 23,351 individuals in iPSYCH2012 and iPSYCH2015i respectively as covariates to control for population structure in each of the cohorts.

**Mendelian Randomization analysis considerations**

If the genetic variants used as instruments are valid, MR can estimate the true causal effect of the exposure on the outcome unbiased from any unobserved confounding factors[15]. We therefore verify that the F statistics for genetic effects, used as instruments to represent for each PHQ9 and WorstEpisode symptom as exposure, are above 20 and therefore strong enough in all analyses (**Supplementary Table 5, 6, 8**). Following the guidelines for MR[16] we adopt a careful interpretation of the MR effect estimates as the association of the genetically predicted levels of the exposure with the outcome.

Further, horizontal pleiotropy is one of the key violations of the MR assumptions[15]. We consequently conducted pleiotropy-robust MR models including Weighted Median MR[17] and MR Egger[18] to compare the MR estimates between different MR models (**Supplementary Table 5, 6, 8**). Each of these methods provides a statistically consistent estimator of the true causal estimate under different assumptions.

**Factor analyses using GenomicSEM**

As genomicSEM[19] works on the genetic covariance matrix, which are not affected by $h^2_{SNP}$ of individual symptoms, we directly compared the genetic covariances of WorstEpisode and PHQ9 symptoms in addition to their rGs. We find that the mean liability scale genetic covariance between all WorstEpisode symptoms is 0.055 (sd = 0.028) while that of PHQ9 symptoms is 0.083 (sd = 0.017), consistent with the higher rGs among PHQ9 symptoms.

To make analysis between PHQ9 and WorstEpisode symptoms comparable in terms of input, we perform the EFA and CFA analysis on PHQ9 both with and without symptom A5 (which is not collected for WorstEpisode symptoms). The results between including and excluding A5 are highly consistent. When A5 is excluded, the two-factor solution ($\chi2$=39.65; AIC=73.65; CFI=0.992; SRMR=0.051) outperforms the one-factor solution ($\chi2$=51.73; AIC=83.73; CFI=0.988; SRMR=0.058), while the three factor model did not converge.

### Supplementary Figures

| Symptom | Data | SNP | OR | LCI | UCI | P value |
|---|---|---|---|---|---|---|
| A2 | ds_WorstEpisode | rs10959576 | 0.94 | 0.92 | 0.97 | 5.66E-05 |
| A2 | PHQ9 | rs10959576 | 0.98 | 0.96 | 1.01 | 2.81E-01 |

| Symptom | Data | SNP | OR | LCI | UCI | P value |
|---|---|---|---|---|---|---|
| A2 | ds_WorstEpisode | rs61975852 | 1.06 | 1.03 | 1.09 | 9.27E-05 |
| A2 | PHQ9 | rs61975852 | 1.04 | 1.01 | 1.07 | 5.99E-03 |

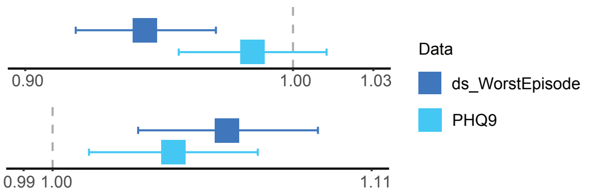


| Symptom | Data | SNP | OR | LCI | UCI | P value |
|---|---|---|---|---|---|---|
| A4 | WorstEpisode | rs11752386 | 1.02 | 0.98 | 1.06 | 3.98E-01 |
| A4 | ds_PHQ9 | rs11752386 | 1.08 | 1.04 | 1.12 | 5.70E-05 |

| Symptom | Data | SNP | OR | LCI | UCI | P value |
|---|---|---|---|---|---|---|
| A4 | WorstEpisode | rs62158170 | 0.99 | 0.96 | 1.03 | 6.88E-01 |
| A4 | ds_PHQ9 | rs62158170 | 0.94 | 0.91 | 0.98 | 1.55E-03 |

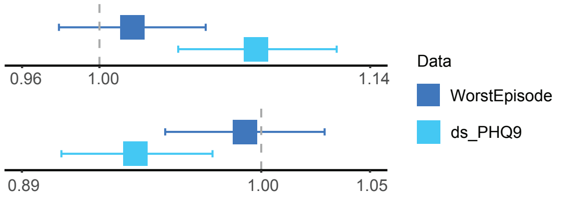


**Supplementary Figure 1**

Forest plots and accompanying data showing the odds ratios (OR) and P-values at significant loci for WorstEpisode or PHQ9 symptoms, in down-sampled data; for each corresponding pair of PHQ9 and WorstEpisode symptom, we downsample the one with higher effective sample sizes accounting for imbalance between cases and controls ($N_{eff} = 4/(1/N_{cases} + 1/N_{controls})$) to the same $N_{eff}$ of the one with lower $N_{eff}$ (**Methods**), keeping its prevalence unchanged. Statistics at the corresponding WorstEpisode or PHQ9 symptoms are shown for comparison; error bars show 95% confidence intervals of the OR estimates.

| Exposure | Outcome | OR | 95% LCI | 95% UCI | P Value |
|---|---|---|---|---|---|
| **Subjective well-being** In general how happy you are? | PHQ9A1 | 0.430 | 0.266 | 0.695 | **5.79E-04** |
| | WorstEpisodeA1 | 0.831 | 0.556 | 1.241 | 3.65E-01 |
| **Insomnia** Do you have trouble falling asleep at night or do you wake up in the middle of the night? | PHQ9A4 | 2.954 | 2.586 | 3.374 | **2.75E-57** |
| | WorstEpisodeA4 | 1.526 | 1.187 | 1.961 | **9.74E-04** |
| | WorstEpisodeA4a (can't fall asleep) | 1.399 | 1.083 | 1.807 | 1.01E-02 |
| | WorstEpisodeA4b (sleeping too much) | 0.806 | 0.616 | 1.055 | 1.16E-01 |
| | WorstEpisodeA4c (waking too early) | 1.322 | 1.040 | 1.680 | 2.24E-02 |
| **Meaning in life** To what extent do you feel your life to be meaningful? | PHQ9A7 | 0.486 | 0.334 | 0.705 | **1.46E-04** |
| | WorstEpisodeA7 | 0.428 | 0.276 | 0.666 | **1.67E-04** |
| **Not worth living** Many people have thoughts that life is not worth living. Have you felt that way? | PHQ9A9 | 2.456 | 2.017 | 2.991 | **3.62E-19** |
| | WorstEpisodeA9 | 1.427 | 1.263 | 1.612 | **1.06E-08** |

**Supplementary Figure 2**

UVMR results where PHQ9 and WorstEpisode symptoms are modeled as outcomes of corresponding traits; significant P-values for PHQ9 symptoms are shown in light blue; significant P-values for WorstEpisode symptoms are shown in darker blue.

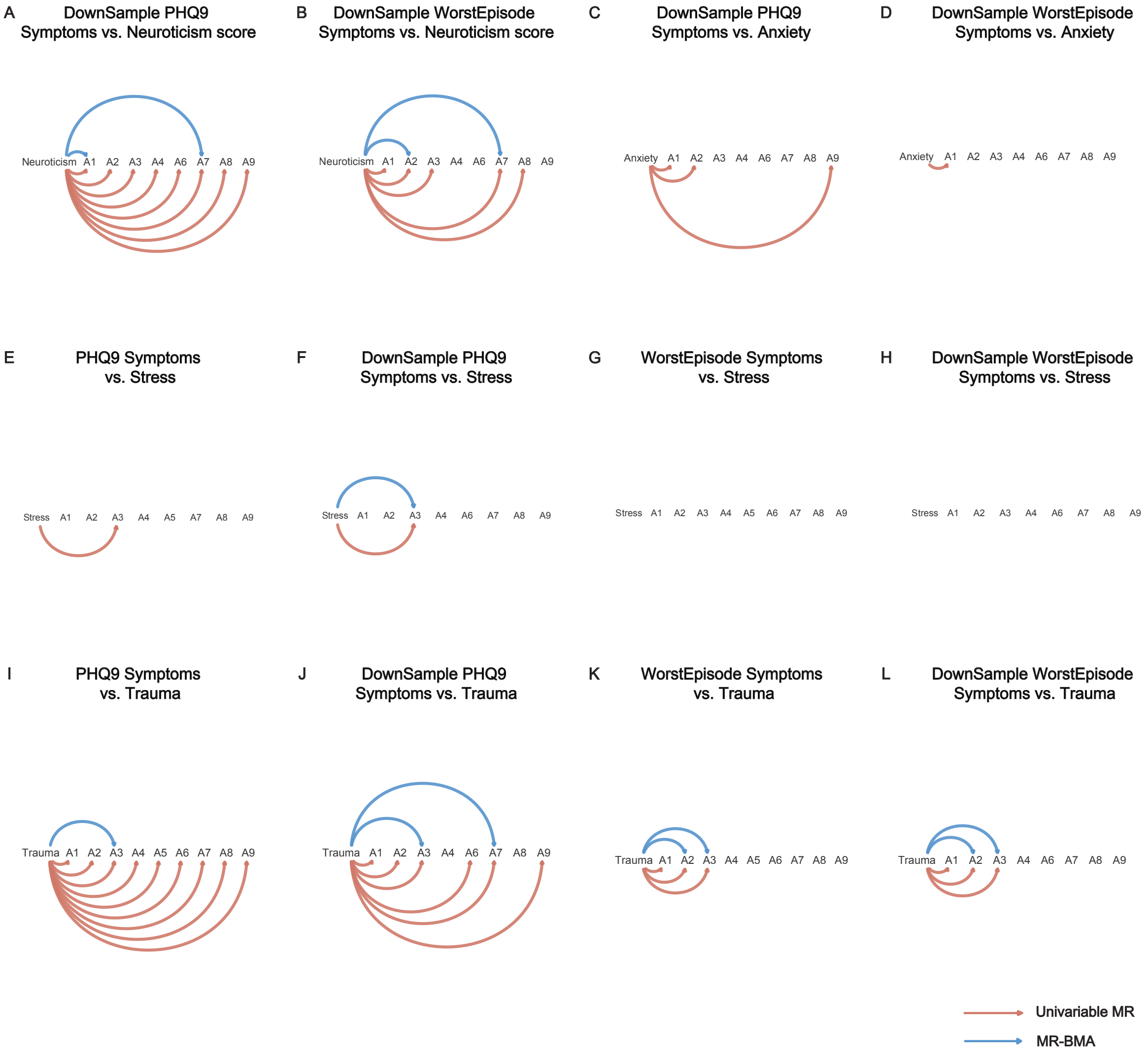


**Supplementary Figure 3**

**A-D.** UVMR results between neuroticism or anxiety (exposures) and downsampled PHQ9 or WorstEpisode symptoms (outcomes) (**Methods**); **E-L**. UVMR results between stress or trauma (exposures) and full-sample or downsampled PHQ9 or WorstEpisode symptoms (outcomes); for all plots, orange arrows indicate significant UVMR results (P<0.05 after Bonferroni correction), blue arrows indicate significant MR-BMA results (<0.05 FDR). Marginal inclusion threshold for MR-BMA is >0.5.

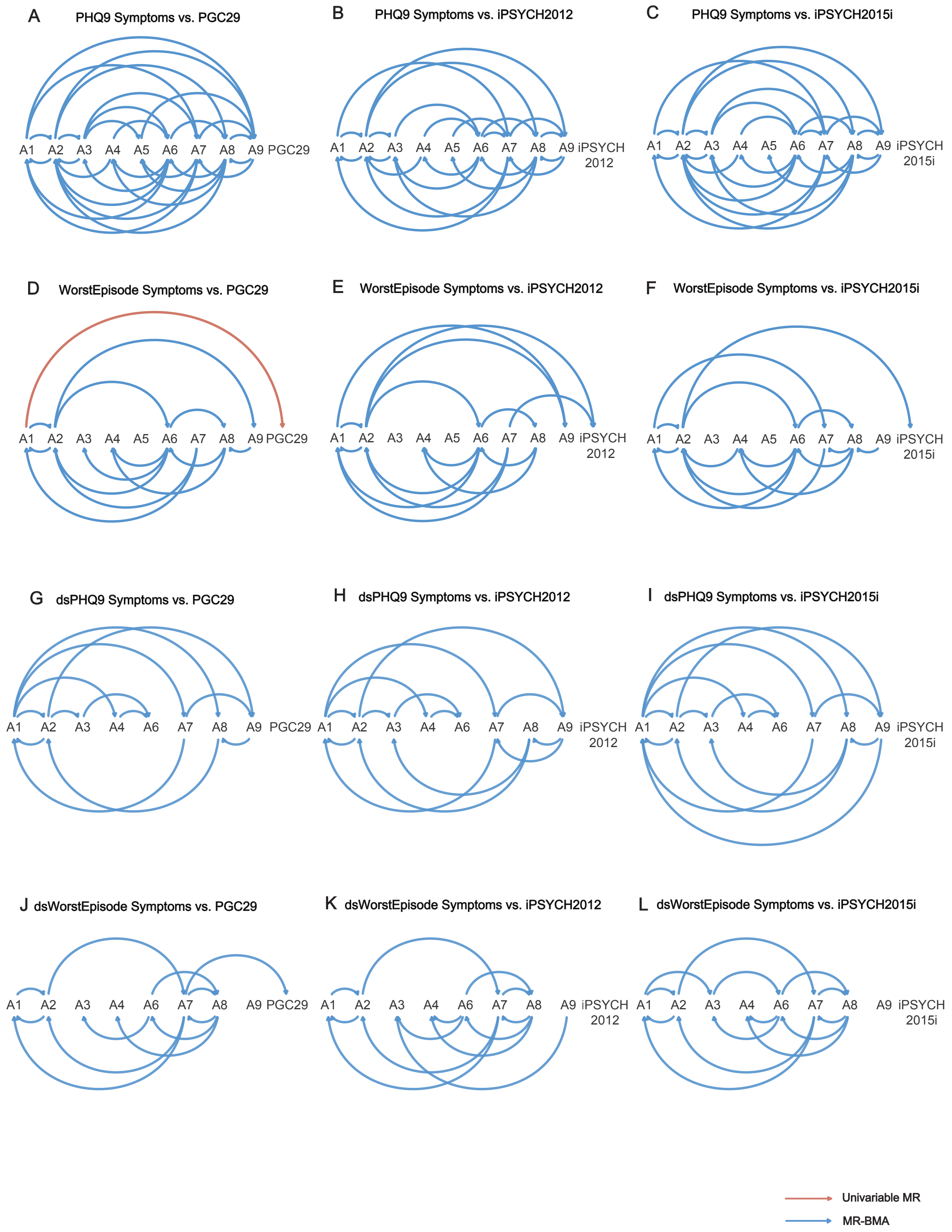
A PHQ9 Symptoms vs. PGC29
B PHQ9 Symptoms vs. iPSYCH2012
C PHQ9 Symptoms vs. iPSYCH2015i
D WorstEpisode Symptoms vs. PGC29
E WorstEpisode Symptoms vs. iPSYCH2012
F WorstEpisode Symptoms vs. iPSYCH2015i
G dsPHQ9 Symptoms vs. PGC29
H dsPHQ9 Symptoms vs. iPSYCH2012
I dsPHQ9 Symptoms vs. iPSYCH2015i
J dsWorstEpisode Symptoms vs. PGC29
K dsWorstEpisode Symptoms vs. iPSYCH2012
L dsWorstEpisode Symptoms vs. iPSYCH2015i
A1 A2 A3 A4 A5 A6 A7 A8 A9 PGC29
iPSYCH 2012
iPSYCH 2015i
Univariable MR
MR-BMA

**Supplementary Figure 4**

**A-F.** UVMR results between PHQ9 or WorstEpisode symptoms and MDD in PGC29[20], iPSYCH2012[3] and iPSYCH2015i[21]; **G-L**. UVMR results between downsampled PHQ9 or WorstEpisode symptoms and MDD in PGC29, iPSYCH2012 and iPSYCH2015i; for all plots, orange arrows indicate significant UVMR results (P<0.05 after Bonferroni correction), blue arrows indicate significant MR-BMA results (<0.05 FDR). Marginal inclusion threshold for MR-BMA is >0.5.

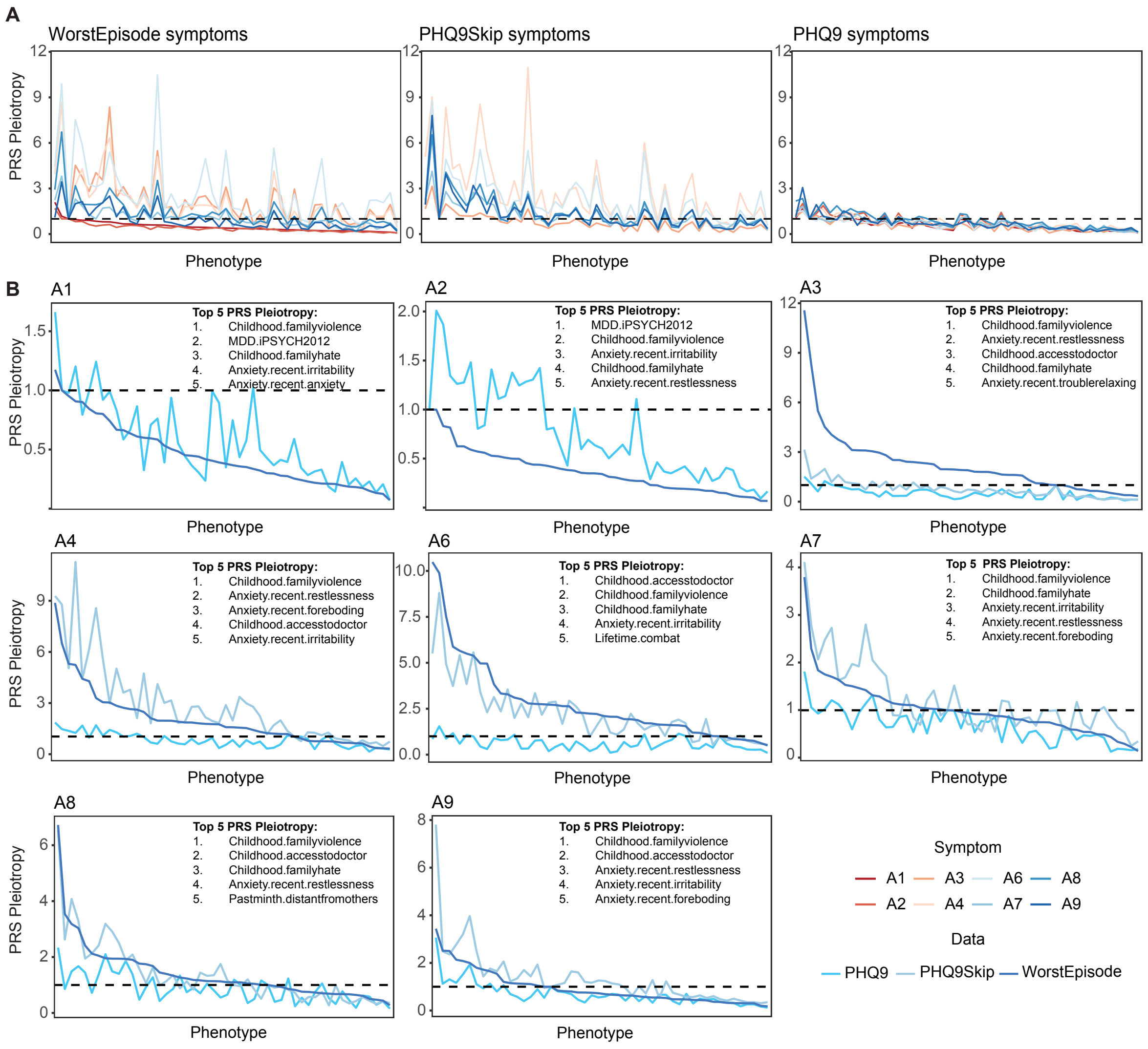


**Supplementary Figure 5**

**A.** Mean PRS Pleiotropy for MDD across 10-fold cross-validation of WorstEpisode, PHQ9 and PHQ9Skip symptoms on 50 phenotypes (including iPSYCH2012 and 49 non-MDD phenotypes, PRS Pleiotropy = $R^2_{non\text{-}MDD}/R^2_{MDD}$); the MDD phenotype here is ICD-10 code defined MDD in iPSYCH2012[3] (**Supplementary Methods**); phenotypes on the x-axis across all three panels are ordered by WorstEpisode A1 PRS Pleiotropy in descending order. **B.** Mean PRS Pleiotropy for each symptom across 10-fold cross validation; phenotypes on the x-axis in each panel are ordered by the WorstEpisode symptom PRS Pleiotropy in descending order; top 5 phenotypes in terms of PRS Pleiotropy are indicated.

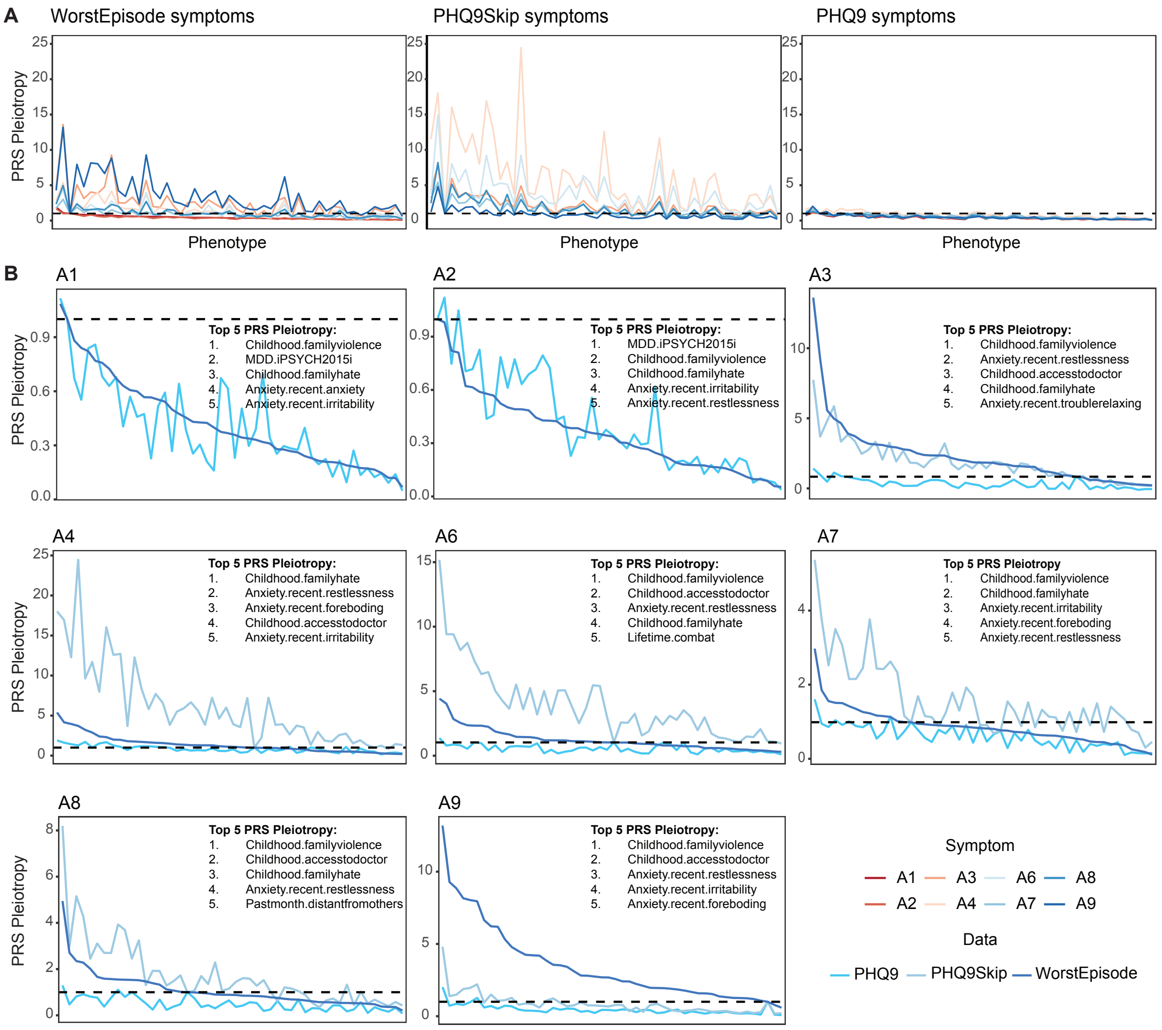


**Supplementary Figure 6**

**A.** Mean PRS Pleiotropy for MDD across 10-fold cross-validation of WorstEpisode, PHQ9 and PHQ9Skip symptoms on 50 phenotypes (including iPSYCH2015i and 49 non-MDD phenotypes, PRS Pleiotropy = $R^2_{non\text{-}MDD}/R^2_{MDD}$); the MDD phenotype here is ICD10 code defined MDD in iPSYCH2015i[21] (Supplementary Methods); phenotypes on the x-axis across all three panels are ordered by WorstEpisode A1 PRS Pleiotropy in descending order. **B.** Mean PRS Pleiotropy for each symptom across 10-fold cross validation; phenotypes on the x-axis in each panel are ordered by the WorstEpisode symptom PRS Pleiotropy in descending order; top 5 phenotypes in terms of PRS Pleiotropy are indicated.

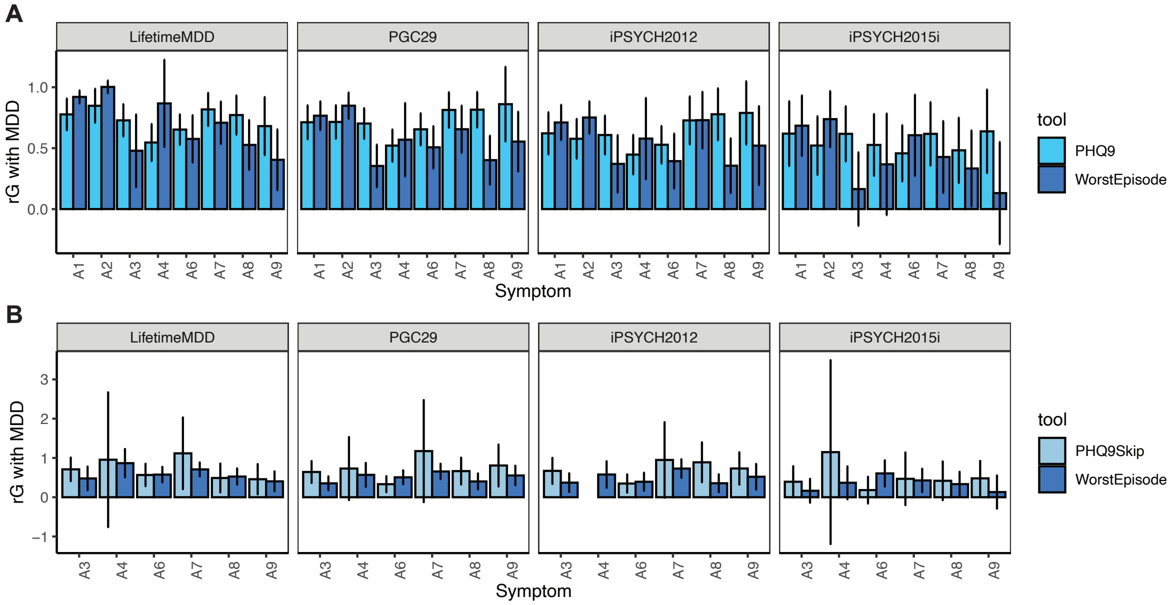


**Supplementary Figure 7**

**A.** Genetic correlations (rG) between WorstEpisode and PHQ9 symptoms A1-A9 with LifetimeMDD[22], and MDD assessed in PGC29[20], iPSYCH2012[3] and iPSYCH2015i[21]; **B.** rG between WorstEpisode and PHQ9Skip symptoms A3-A9 with MDD in all four cohorts.

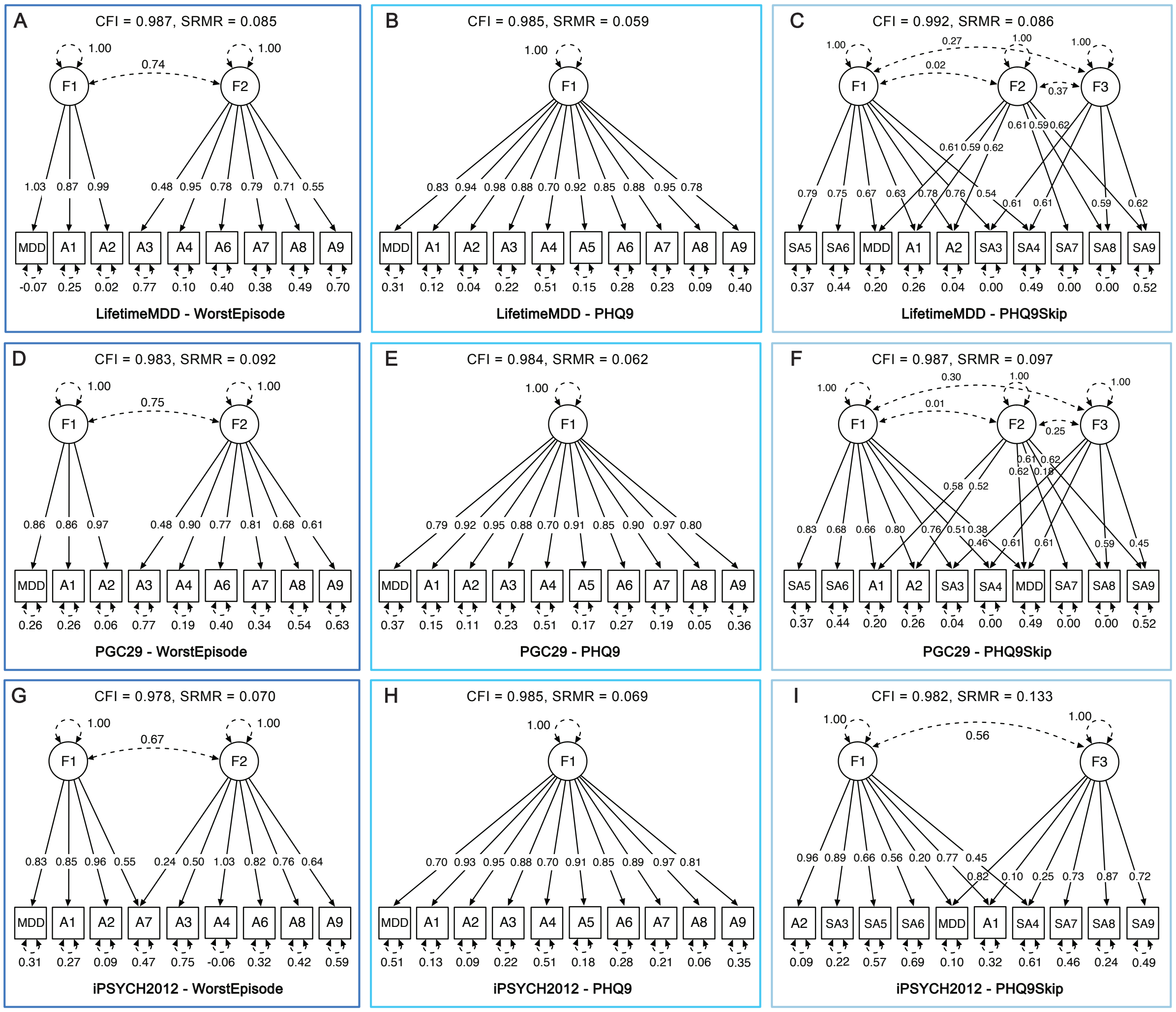


**Supplementary Figure 8**

**A-C.** CFA factor loadings on LifetimeMDD and WorstEpisode, PHQ9 and PHQ9Skip symptoms, excluding symptom A5; **D-F.** CFA factor loadings on ICD10 based MDD defined in iPSYCH2012 and WorstEpisode, PHQ9 and PHQ9Skip symptoms, excluding symptom A5; **H-J.** CFA factor loadings on ICD10 based MDD defined in iPSYCH2015i and WorstEpisode, PHQ9 and PHQ9Skip symptoms, excluding symptom A5; only models with P < 0.05 are considered in model comparisons; models with best fit are shown.

**Supplementary Table Legends**

**Supplementary Table 1.**
**Definition and sample size for WorstEpisode and downsampled WorstEpisode symptoms.** This table shows the definitions and sample sizes of all WorstEpisode symptoms, in a) all individuals who endorsed them, b) only those with LifetimeMDD disease status, and c) when down-sampled to the effective sample size of PHQ9 symptoms (for A1, A2 and A9).

**Supplementary Table 2.**
**Definition and sample size for PHQ9, PHQ9Strict, PHQ9Skip and downsampled PHQ9 symptoms.** This table shows the definition of all PHQ9, PHQ9Strict, PHQ9Skip and downsampled PHQ9 symptoms, where applicable in a) all individuals who endorsed them, b) only those with LifetimeMDD disease status, and c) down-sampled to the effective sample size of WorstEpisode symptoms(for A3, A4, A6, A7 and A8).

**Supplementary Table 3.**
**Risk alleles that are genome-wide significantly associated with either WorstEpisode or PHQ9 symptoms.** OR: odds ratio. LCI: the lower 95% confidence interval. UCI: the upper 95% confidence interval. Symptom A3a is not assessed in PHQ9.

**Supplementary Table 4.**
**Definition and sample size for non-MDD phenotypes.** This table shows the definition of all non-MDD phenotypes used in this manuscript, including individual items of neuroticism, anxiety, stress and trauma.

**Supplementary Table 5.**
**Univariable Mendelian Randomization (UVMR) results where PHQ9 and WorstEpisode symptoms are modeled as outcomes of corresponding traits.** Methods including Inverse-Variance Weighted (IVW), Weighted Median (WM) and MR-Egger are used. F stats: F statistics; n_SNP: number of SNPs acting as instruments; se: standard error; lci: the lower 95% confidence interval; uci: the upper 95% confidence interval; bf: Bonferroni-corrected p values.

**Supplementary Table 6.**
**Univariable Mendelian Randomization (UVMR) results between MDD symptoms and non-MDD phenotypes.** Methods including Inverse-Variance Weighted (IVW), Weighted Median (WM) and MR-Egger are used. F stats: F statistics; n_SNP: number of SNPs acting as instruments; se: standard error; lci: the lower 95% confidence interval; uci: the upper 95% confidence interval; bf: Bonferroni-corrected p values.

**Supplementary Table 7.**
**Two-sample multivariable MR based on Bayesian model averaging (MR-BMA) results between MDD symptoms and non-MDD phenotypes.** MR-BMA results showing effects of neuroticism, anxiety, stress or trauma on PHQ9, downsampled PHQ9, WorstEpisode and downsampled WorstEpisode symptoms.

**Supplementary Table 8.**

**Univariable Mendelian Randomization (UVMR) results between MDD symptoms and MDD from replication cohorts.** Methods including Inverse-Variance Weighted (IVW), Weighted Median (WM) and MR-Egger are used. F stats: F statistics; n_SNP: number of SNPs acting as instruments; se: standard error; lci: the lower 95% confidence interval; uci: the upper 95% confidence interval; bf: Bonferroni-corrected p values.

**Supplementary Table 9.**

**Two-sample multivariable MR based on Bayesian model averaging (MR-BMA) results between MDD symptoms and MDD from replication cohorts.** MR-BMA results showing effects of MDD in PGC29, iPSYCH2012 and iPSYCH2015i on PHQ9, downsampled PHQ9, WorstEpisode and downsampled WorstEpisode symptoms. Only results that have marginal inclusion >0.5 and FDR-corrected P value <0.05 are shown in this table.

**Supplementary Table 10-12.**

**PRS Pleiotropy of MDD symptoms on 50 non-MDD phenotypes.** Mean PRS R2s on 50 non-MDD phenotypes using 10-fold cross-validated PRS from PHQ9, PHQ9Skip and WorstEpisode symptoms, and PRS Pleiotropy calculated using their ratio with the corresponding PRS R2s (PRSPleiotropy = $R^2_{\text{non-MDD}}/R^2_{\text{MDD}}$) on LifetimeMDD in UKBiobank (**Supplementary Table 10**) or ICD10-based MDD in iPSYCH2012 (**Supplementary Table 11**) and iPSYCH2015i (**Supplementary Table 12**).

**Supplementary References**


1. Bycroft, C. *et al.* The UK Biobank resource with deep phenotyping and genomic data. *Nature* **562**, 203–209 (2018).
2. Abraham, G., Qiu, Y. & Inouye, M. FlashPCA2: principal component analysis of Biobank-scale genotype datasets. *Bioinformatics* **33**, 2776–2778 (2017).
3. Pedersen, C. B. *et al.* The iPSYCH2012 case-cohort sample: new directions for unravelling genetic and environmental architectures of severe mental disorders. *Mol. Psychiatry* **23**, 6–14 (2018).
4. Bybjerg-Grauholm, J. *et al.* The iPSYCH2015 Case-Cohort sample: updated directions for unravelling genetic and environmental architectures of severe mental disorders. Preprint at https://doi.org/10.1101/2020.11.30.20237768.
5. Nørgaard-Pedersen, B. & Hougaard, D. M. Storage policies and use of the Danish Newborn Screening Biobank. *J. Inherit. Metab. Dis.* **30**, 530–536 (2007).
6. Mors, O., Perto, G. P. & Mortensen, P. B. The Danish Psychiatric Central Research Register. *Scand. J. Public Health* **39**, 54–57 (2011).
7. Lynge, E., Sandegaard, J. L. & Rebolj, M. The Danish National Patient Register. *Scand. J. Public Health* **39**, 30–33 (2011).
8. Pedersen, C. B. The Danish Civil Registration System. *Scand. J. Public Health* **39**, 22–25 (2011).
9. Browning, B. L., Zhou, Y. & Browning, S. R. A One-Penny Imputed Genome from Next-Generation Reference Panels. *Am. J. Hum. Genet.* **103**, 338–348 (2018).
10. Browning, S. R. & Browning, B. L. Rapid and accurate haplotype phasing and missing-data inference for whole-genome association studies by use of localized haplotype clustering. *Am. J. Hum. Genet.* **81**, 1084–1097 (2007).
11. McCarthy, S. *et al.* A reference panel of 64,976 haplotypes for genotype imputation. *Nat. Genet.* **48**, 1279–1283 (2016).

12. Manichaikul, A. *et al.* Robust relationship inference in genome-wide association studies. *Bioinformatics* **26**, 2867–2873 (2010).
13. Price, A. L. *et al.* Principal components analysis corrects for stratification in genome-wide association studies. *Nat. Genet.* **38**, 904–909 (2006).
14. Choi, S. W. & O'Reilly, P. F. PRSice-2: Polygenic Risk Score software for biobank-scale data. *Gigascience* **8**, (2019).
15. Burgess, S., Foley, C. N. & Zuber, V. Inferring Causal Relationships Between Risk Factors and Outcomes from Genome-Wide Association Study Data. *Annu. Rev. Genomics Hum. Genet.* **19**, 303–327 (2018).
16. Burgess, S. *et al.* Guidelines for performing Mendelian randomization investigations. *Wellcome Open Res* **4**, 186 (2019).
17. Bowden, J., Davey Smith, G., Haycock, P. C. & Burgess, S. Consistent Estimation in Mendelian Randomization with Some Invalid Instruments Using a Weighted Median Estimator. *Genet. Epidemiol.* **40**, 304–314 (2016).
18. Bowden, J., Davey Smith, G. & Burgess, S. Mendelian randomization with invalid instruments: effect estimation and bias detection through Egger regression. *Int. J. Epidemiol.* **44**, 512–525 (2015).
19. Grotzinger, A. D. *et al.* Genomic structural equation modelling provides insights into the multivariate genetic architecture of complex traits. *Nat Hum Behav* **3**, 513–525 (2019).
20. Wray, N. R. *et al.* Genome-wide association analyses identify 44 risk variants and refine the genetic architecture of major depression. *Nat. Genet.* **50**, 668–681 (2018).
21. Bybjerg-Grauholm, J. *et al.* The iPSYCH2015 Case-Cohort sample: updated directions for unravelling genetic and environmental architectures of severe mental disorders. *bioRxiv* (2020) doi:10.1101/2020.11.30.20237768.
22. Cai, N. *et al.* Minimal phenotyping yields genome-wide association signals of low specificity for major depression. *Nat. Genet.* **52**, 437–447 (2020).
